# Physical fitness supports brain maintenance and cognitive reserve in cognitively normal aging: a cross-sectional examination

**DOI:** 10.64898/2026.01.14.26344098

**Authors:** Svenja Schwarck, Niklas Behrenbruch, Beate Schumann-Werner, Eóin N. Molloy, Berta Garcia-Garcia, Anne Hochkeppler, Larissa Fischer, Anna-Therese Büchel, Enise I. Incesoy, Jose Bernal, Roberto Duarte Coello, Maria d.C. Valdés-Hernández, Joanna M. Wardlaw, Niklas Vockert, Patrick Müller, Gusalija Behnisch, Barbara Morgado, Hermann Esselmann, Constanze I. Seidenbecher, Björn H. Schott, Henryk Barthel, Osama Sabri, Jens Wiltfang, Michael C. Kreissl, Emrah Düzel, Anne Maass

**Author notes:** Corresponding author: Anne Maass, Ph.D., Email Adress.

## Abstract

**Background:** Preserving cognitive and brain health is central for healthy aging. Cognitive reserve (CR) and brain maintenance (BM) support resilience against age- and disease-related cognitive decline. Physical fitness represents a plausible pathway to both CR and BM, yet the underlying neurobiological mechanisms remain insufficiently understood. This study examined how fitness relates to Alzheimer’s pathology and whether it moderates or mediates the pathology–cognition relationship.

**Methods:** Data were obtained from 345 cognitively unimpaired older adults (mean age = 73.11 ± 8.03 years; 177 females) participating in the ongoing SFB1436 study. We assessed global cognition and delayed verbal memory performance, aerobic fitness (VO₂max), and muscular capacity. Blood biomarkers included plasma Aβ₁₋₄₂/Aβ₁₋₄₀, p-tau217, and GFAP (glial fibrillary acidic protein), and serum BDNF (brain-derived neurotrophic factor), VEGF (vascular endothelial growth factor), and Cathepsin-B. Neuroimaging measures comprised medial temporal lobe (MTL) tau burden ([¹⁸F]PI-2620 PET), MRI-derived hippocampal volumes, white matter hyperintensities, perivascular spaces (PVS) in basal ganglia (BG) and centrum semiovale, gray matter volume (GMV), and MTL thickness.

To examine BM, we tested associations between fitness and brain pathology. To examine CR, we conducted moderation analyses assessing whether fitness attenuated the negative impact of pathology on cognition. Mediation analyses further evaluated whether hippocampal volume, total GMV, or MTL thickness mediated a potential association between fitness and cognition. All models controlled for age and sex (and education in mediation analyses), and multiple comparisons were FDR-corrected.

**Results:** Physical fitness was not related to cognitive performance. Higher muscular capacity was associated with lower BG PVS volumes, while higher aerobic fitness was related to higher MTL thickness and total GMV. Elevated MTL tau burden was associated with poorer verbal memory performance. Although physical fitness did not significantly moderate the tau–memory relationship, model comparison provided weak evidence for CR effects (p = .015).

**Conclusion:** Muscular capacity was linked to lower BG PVS volumes, supporting resistance against pathology, while aerobic fitness related to preserved cortical integrity and tended to act as a CR proxy against MTL tau pathology. Physical fitness may support cerebrovascular and glymphatic function, thereby promoting cognitive resilience and extending healthspan by sustaining functional brain health and mitigating tau-related cognitive decline.

**Trial Registration:** The study was retrospectively registered in the German Clinical Trials Register (DRKS00032449; date of registration: 2025-05-07; recruiting ongoing).

## Background

Aging constitutes a multidimensional developmental process associated with gradual cognitive decline and increased susceptibility for metabolic, cardiovascular, cerebrovascular, and neurodegenerative diseases, including Alzheimer’s disease (AD) [1–6]. Yet, the rate and magnitude of cognitive decline varies widely [1,7], with some individuals maintain high cognitive performance despite age or disease, while other decline without detectable pathology [4,8,9]. Research has focused on factors promoting resilience, defined as the brain’s ability to preserve cognition and function despite aging or diseases-related pathology [10,11]. Among these factors, physical activity and structured exercise have emerged as promising contributors to preserved cognition, neuroplasticity, and brain health in older adults [12–14],. These effects may occur through mechanisms involving cognitive reserve (CR) and brain maintenance (BM), core constructs of resilience that support healthy aging and the extension of healthspan [10,11].

Brain maintenance refers to the capacity to preserve cognitive function in older age by resisting the decline of neural resources and the onset of neuropathological alterations [10,15]. Typical age-related changes in the brain include the accumulation of Alzheimer’s disease-related proteins, white matter lesions, enlargement of perivascular spaces (ePVS) and atrophy of gray matter [10,16–19]. The medial temporal lobe is especially vulnerable for age-related atrophy and early accumulation of tau tangles [20–22] Although longitudinal studies best capture trajectories of BM [10,11], cross-sectional designs also provide valuable insights into the associations between lifestyle and neuropathology. In this respect, physical activity has repeatedly been linked to structural brain health, such as preserved gray matter volume and white matter integrity [14,23–25]. Yet, the relationship to age-related pathological changes such as ePVS, a potential marker for impaired glymphatic clearance [26–28], or to the accumulation of tau tangles in the medial temporal lobe (MTL) remain unclear. Some previous studies reported no association between physical activity and AD markers, such as plasma Aβ₁₋₄₂/ Aβ₁₋₄₀ or Aβ PET burden in older adults with mild cognitive impairment or cognitively unimpaired older adults [29–32]. On the other hand, recent studies found a link between higher physical activity and lower plasma p-tau217 levels [33] and slower changes in p-tau181 concentrations over time [34]. Higher physical activity has been further linked to lower Flortaucipir tau PET burden, both in a small study using self-reported activity [35] and in another study using step counts of seven consecutive days [36]. However, the association between MTL tau burden and objective physical fitness measures, such as aerobic fitness, remains underexplored.

While BM denotes resistance to pathological changes, cognitive reserve, a central component of resilience, denotes the brain’s capacity to compensate for age-related changes or neuropathological damage [10,11,15]. This is shaped through diverse life experiences [10,37]. Frequently operationalized through socio-behavioral proxies such as education, CR is also modulated by lifestyle factors and may explain variability in cognitive aging trajectories [10,37,38]. Evidence supporting physical activity as a contributor to CR remains mixed, ranging from positive effects [39–42], to null findings [43–45]. These inconsistencies likely reflect differences in the pathology markers, the measurement of physical activity, potential cohort biases, and statistical approaches that were used [10,11,38], even in the presence of common guidelines.

Epidemiological studies consistently report positive associations between sustained physical activity and cognitive performance in older adults, including preserved gray matter integrity and increased secretion of neurotrophic factors [12,14,46]. Physical activity has been linked to larger hippocampal volumes across the lifespan, with some evidence highlighting effects in the left anterior hippocampus [47–51]. Neuroplasticity-related factors, such as BDNF, VEGF, and Cathepsin B, are implicated in this relationship due to their roles in synaptic modulation and neuronal survival [52–56]. Chronic physical exercise enhances myokine release [57,58], i.e., muscle contractions trigger the secretion of myokines such as irisin and Cathepsin B, which can cross the blood-brain barrier and seem to stimulate neurotrophic factors like BDNF, supporting neurogenesis [57,59–61]. Peripheral levels of BDNF correlate positively with physical activity, hippocampal volume, and cognitive performance in older adults [12,13,47,62,63]. Although Cathepsin B contributes to angiogenesis and neurogenesis, its direct association with physical activity and cognitive performance, as well as its potential mediation of exercise-related cognitive effects, remain inconsistent [64–67]. Additionally, anaerobic exercise elevates muscle-derived lactate, which seems to cross the blood-brain-barrier, activates hydroxycarboxylic acid receptor 1 (HCAR1), upregulates VEGF, and promotes angiogenesis, potentially enhancing cognitive function [68,69].

Overall, brain structural integrity, particularly hippocampal volume, appears to mediate the relationship between physical activity and cognitive performance [24,50,70]. Nevertheless, the underlying neurobiological mechanisms remain poorly understood, and only a limited number of cross-sectional studies have identified hippocampal volume as a mediator [14,24,47].

In summary, accumulating evidence suggests a link between physical activity and both cognitive performance and brain integrity [12,14,47], as well as the role of physical activity in promoting resilience, including CR [39–42] and BM [14,23,24,33–36]. However, most cross-sectional studies rely on self-reported physical activity questionnaires, which are vulnerable to recall bias and often fail to capture actual activity levels [71]. Objective measures such as aerobic (VO₂max, maximal oxygen uptake) and muscular fitness (as the inverse of sarcopenia), provide standardized indices of physical fitness [72]. Both measures have been associated with cognitive performance before [13,14,24,73,74], offering complementary perspectives on inter-individual variability in cognitive outcomes.

To address these limitations, the present study implemented a comprehensive assessment, comprising objective physical fitness measures, deep neuropsychological phenotyping, blood-based biomarkers related to AD pathology and plasticity, objective physical fitness measures, and imaging-based brain and pathology outcomes in a cognitively normal aging cohort. Regional tau burden in the MTL was assessed using the second-generation tau tracer [18F]PI-2620. We addressed four key questions: (1) whether higher physical fitness - both aerobic and muscular - is associated with better cognitive performance; (2) whether physical fitness is linked to lower brain pathology, supporting the concept of brain maintenance; (3) whether physical fitness serves as a proxy for cognitive reserve; and (4) whether hippocampal volume - especially in the left anterior region - and neuroplasticity markers like BDNF mediate the relationship between physical fitness and cognitive outcomes.

## Methods

This study analyzed baseline data from an ongoing cohort investigating cognitively healthy older adults (for details see [75]). The cohort is part of the central project *“Z03 – Human Molecular Imaging of Aging and SuperAging”* within the Collaborative Research Centre 1436, *“Neural Resources of Cognition”*. Its aim is to characterize factors or mechanisms related to successful brain and cognitive aging in deeply phenotyped older individuals using a combination of cross-sectional and longitudinal assessments. Participants undergo a comprehensive evaluation including neuropsychological testing, structural and functional neuroimaging, tau PET, molecular and genetic biomarker analyses, physical fitness assessments, and lifestyle questionnaires. For the present study, only baseline data were used.

### Participants

The sample comprised 345 cognitively normal older adults (177 females), aged 60– 94 years (M = 72.77, SD = 7.95), all independently living and with relatively high educational attainment (M = 14.93 years, SD = 2.38). Normal cognition was determined using the CERAD-Plus normative scores adjusted for age, sex, and education (more details give below).

Exclusion criteria were any presence of psychiatric or neurological disorders, major systemic illnesses, diagnosed diabetes, medications affecting the central nervous system (e.g., antidepressants, hypnotics, certain analgesics), and cardiovascular, musculoskeletal, endocrine, pulmonary, or neurodegenerative conditions. Recruitment was conducted via public outreach, including media campaigns and targeted mailings coordinated with local authorities.

The study design and protocol were approved by the Ethics Committee of the Medical Faculty at Otto-von-Guericke University Magdeburg (approval number 200/19) and conducted in accordance with the code of ethics of the World Medical Association (Declaration of Helsinki, 1967). All participants provided written informed consent and received financial compensation for participation. Study registration to the “German Clinical Trials Register” https://drks.de has been performed and the registration is currently processed (number: DRKS00032449).

### Experimental design

All analyses were based on the baseline data (Fig 1). The protocol included a multimodal assessment comprising: comprehensive neuropsychological testing, high-resolution MRI, [^18^F]PI-2620 tau PET imaging (subsample of participants), venous blood collection for molecular and genetic markers, and objective measures of aerobic and muscular fitness. Testing sessions were distributed to optimize data quality and reduce participant fatigue, ensuring reliable measures across all modalities. The number of available data for each measurement modality is also provided in Figure 1.

**Fig 1.**
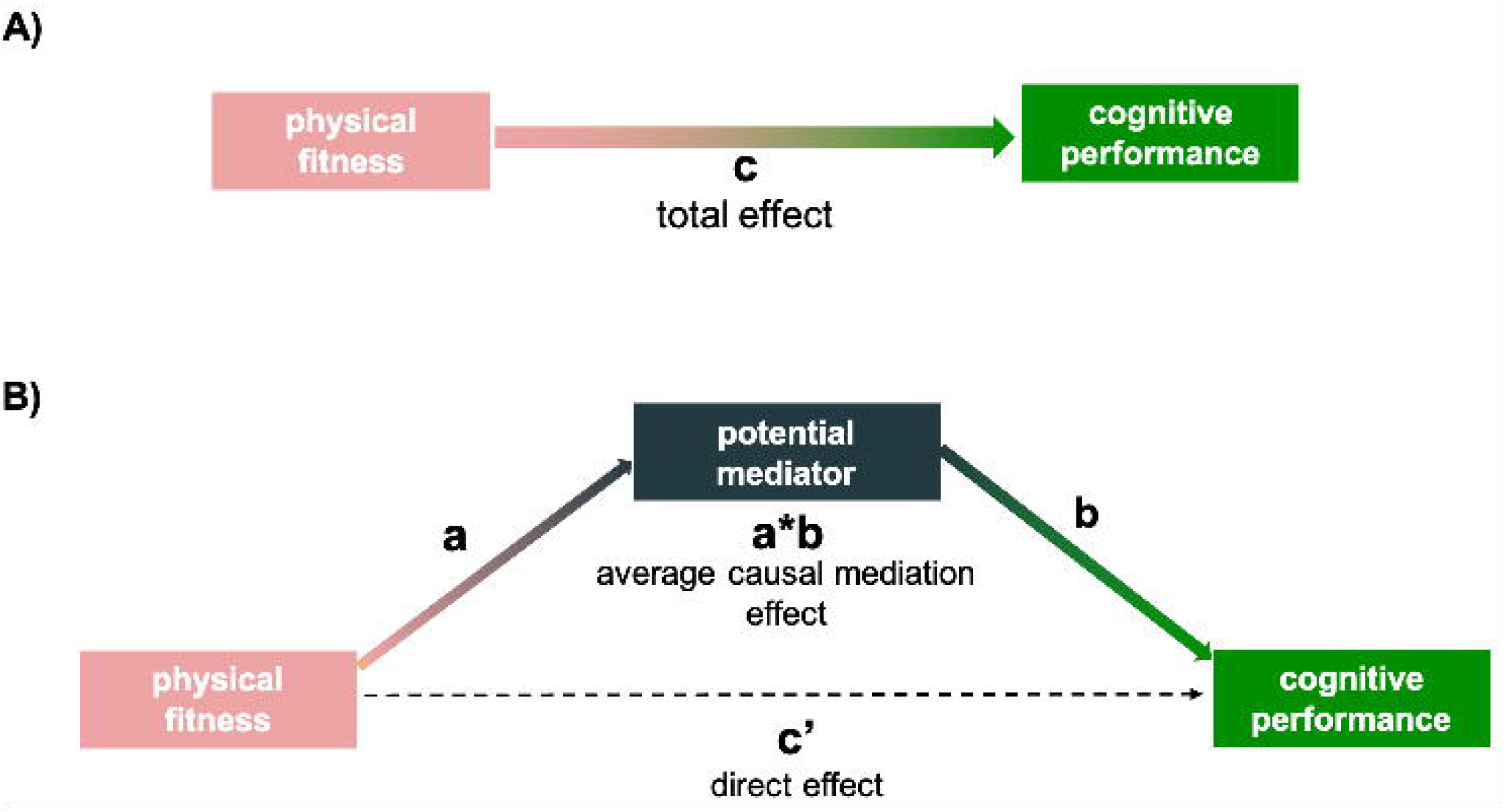
Overview of study methodology and schematic presentation of the primary variables and measures. CERAD, Consortium to establish a Registry for Alzheimer’s Disease (z-composite using CERAD-Plus battery); PACC5, preclinical Alzheimer’s composite (z-composite); VLMT, German adaptation of Rey’s Auditory Verbal Learning Test (delayed verbal memory performance); ROCFT, Rey-Osterrieth Complex Figure Test (visuospatial delayed memory performance); BG PVS, basal ganglia perivascular spaces; CSO PVS, centrum semiovale perivascular spaces; BDNF, brain derived neurotrophic factor; VEGF, vascular endothelial growth factor, GFAP, glial fibrillary acidic protein; VO_2_max, maximal oxygen consumption (ml/min/kg); ASMM, appendicular skeletal muscle mass (in kg); TUG, timed-up-and-go-test (in seconds).

### Neuropsychological measurements

Neuropsychological testing was conducted across two separate sessions. During the initial session, participants (n = 357) underwent the CERAD-Plus battery (Consortium to Establish a Registry for Alzheimer’s Disease; [76]) to confirm normal cognitive performance. Raw scores were converted to age-, sex-, and education-adjusted z-scores. Participants were excluded if scores indicated evidence of possible cognitive limitations based on the neuropsychological Jak/Bondi criteria ([77]) or MMSE < 26 (for details see [75]).

The second testing session occurred approximately one week later and consisted of an extended cognitive battery covering a broader range of domains. Episodic memory was probed using the Verbal Learning and Memory Test (VLMT; [78]), the German analog of the Rey Auditory Verbal Learning Test. Additional memory assessments included the Logical Memory II subtest from the Wechsler Memory Scale (WMS; [79]) and the Free and Cued Selective Reminding Test (FCSRT; [80]). Visuospatial and constructional abilities were examined using the Rey-Osterrieth Complex Figure Test (ROCFT; [81]). Attention and processing speed were evaluated using the Symbol Digit Modalities Test (SDMT; [82]). Verbal fluency was further assessed using both semantic and phonemic fluency tasks, including task-switching components, based on the Regensburger Wortflüssigkeitstest (RWT; [83]), which is comparable to the Controlled Oral Word Association Test (COWAT). A composite z-score was computed following the Preclinical Alzheimer’s Cognitive Composite (PACC5) framework [84] derived by the mean of MMSE, delayed logical memory (WMS-R), SDMT, FCSRT (free and total recall), and RWT category fluency tasks.

### MRI acquisition and processing

High-resolution structural neuroimaging was conducted using a 3 Tesla MRI scanner (MAGNETOM Skyra, Siemens), employing a multi-echo magnetization-prepared rapid acquisition gradient echo (MEMPRAGE) sequence. Images were acquired at an isotropic voxel resolution of 0.8 mm, with echo times (TEs) of 1.85, 3.75, 5.65, and 7.55 ms, repetition time (TR) of 2560 ms, inversion time (TI) of 1100 ms, and a flip angle of 7°. The total scan duration was approximately 6.5 minutes. All subsequent volumetric analyses were conducted on the echo-combined T1-weighted images. Additionally, a Fluid-Attenuated Inversion Recovery (FLAIR) sequence was acquired with 1.0 mm isotropic resolution (TR = 5 s, TE = 393 ms, TI = 1.8 s), resulting in a total acquisition time of 4 min 37 s.

Quantification of hippocampal subfields was performed using the Automatic Segmentation of Hippocampal Subfields (ASHS) algorithm. T1-weighted scans (n = 260) were resampled to a voxel resolution of 0.8 × 0.4 × 0.4 mm, and segmentation was guided by the 3T Pennsylvania Memory Center atlas [85]. This process enabled extraction of volumetric measures for the anterior hippocampus (aHCV), posterior hippocampus (pHCV), and total hippocampal volume (wHCV). The segmentation protocol adhered to the Harmonized Protocol on T1-weighted images [85,86], supplemented by inclusion of the alveus and fimbria regions [87].

Each segmented image was subjected to independent visual inspection by two blinded raters to ensure anatomical accuracy. Manual corrections were applied in 62 hemispheres, addressing systematic segmentation errors such as underestimation of the lateral hippocampal boundary or misclassification of the choroid plexus [85]. Adjustments followed guidelines from the ASHS-T2 and Harmonized Protocols [27,85,88]. Following visual quality control and manual refinement, volumetric data for aHCV, pHCV, and wHCV were extracted for each hemisphere and aggregated to generate bilateral measures for subsequent analyses.

To quantify volumetric brain parameters, high-resolution T1-weighted structural images (n = 263) were processed using Freesurfer to extract total gray matter volume (GMV) and via samseg to extract total intracranial volume (TIV).

To control for interindividual variability in cranial size, a linear regression-based normalization procedure was employed in accordance with established protocols [89]. Specifically, adjusted volumes (Voladj) were computed using the equation:

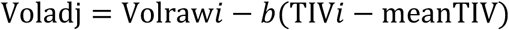

Here, Volrawᵢ represents the unadjusted volume of a given region of interest (ROI) for subject *i*, *b* denotes the slope of the linear regression of raw volume on TIV, and meanTIV corresponds to the sample mean [89]. These adjusted volumetric measures were subsequently used in statistical analyses.

Perivascular spaces (PVS), indicative of cerebral small vessel disease, were segmented within the centrum semiovale (CSO) and basal ganglia (BG) using a validated semi-automated pipeline that integrated information from T1-weighted and FLAIR sequences (n = 237) [27,88]. The CSO ROI encompassed the supratentorial white matter excluding BG structures, while the BG ROI comprised the caudate, lentiform nucleus, thalamus, and adjacent internal and external capsules. Although these regions do not precisely correspond to anatomical boundaries, their delineation follows established conventions from visual rating protocols [90]. PVS burden was quantified as fractional volume (PVS_volume / ROI_volume) and normalized using the bestNormalize algorithm to account for distributional skewness.

White matter hyperintensities (WMH) were segmented from T1-weighted and FLAIR images using the AI-enhanced Lesion Segmentation Toolbox (LST-AI), incorporating deep learning-based lesion detection (n = 237) [91]. Total and regional WMH volumes (cortical lobes, corpus callosum, and internal/external capsules) were quantified based on the USCLobes Atlas [92]. To address non-normality, WMH volumes underwent Box-Cox transformation. TIV correction was achieved via linear regression, with residual values from the TIV-WMH model employed for subsequent analysis.

Medial temporal lobe (MTL) thickness was used from the T1-weighted images using Freesurfer (n = 263). Bilateral MTL thickness (in mm) was calculated as the average across both hemispheres including the MTL regions entorhinal thickness, parahippocampal thickness and fusiform thickness.

### PET acquisition and analysis

Regional tau burden was measured with the second generation tau tracer [18F]PI-2620, selected for its high specificity to tau isoforms relevant to Alzheimer’s disease and reduced off-target binding compared to earlier tracers [93–95]. Simultaneous PET-MRI acquisition was conducted using the Siemens Biograph mMR system at the DZNE in Magdeburg. Participants (n = 143; available scans included until 02/2025) received an intravenous bolus injection of a single average tracer dose of 183 MBq (SD = 1.7 MBq)[93–95]. Dynamic PET data were acquired over 60 minutes and reconstructed into 34 frames: 12×15 s, 6×30 s, 4×60 s, 4×120 s, 4×180 s, 2×300 s, and 2×600 s.

Motion correction and co-registration of PET frames to individual T1-weighted MRIs were performed. T1-weighted images aquired during the MR-PET scan were segmented with FreeSurfer to derive regions-of-interest.^1^ Distribution volume ratio (DVR) maps were computed using the Multilinear Reference Tissue Model 2 (MRTM2) implemented in QModeling (Matlab R2024V) [96,97]. The globus pallidus and inferior cerebellum were designated as high-binding and reference regions, respectively [94,98]. Mean DVR values were extracted for key MTL structures, including the entorhinal cortex, parahippocampal gyrus, fusiform gyrus, and amygdala.

### Blood-Based Biomarkers

On the same day as the physical fitness assessment (see below), venous blood was collected under standardized fasting and substance-abstention conditions (2 h for food, 4 h for nicotine/caffeine). Serum samples were cooled, centrifuged (15 min, 4°C, 1100 × g), aliquoted, and stored at −80°C. Target analytes were quantified via enzyme-linked immunosorbent assays (ELISA) in duplicate: BDNF (custom sandwich ELISA adapted from [99]; n = 89), VEGF (R&D Systems; n = 86), and Cathepsin B (Abcam; n = 87). Analyses adhered to quality guidelines from the FDA (2001) and EMA (2013). On a separate occasion, EDTA plasma samples were obtained. Blood biomarkers for amyloid and tau pathology were analyzed at the department of Psychiatry and Psychotherapy, University Medical Center Goettingen & DZNE, Goettingen. Aliquots of 500 μl EDTA–blood plasma stored at −80 °C in Matrix 0.5 mL tubes (Thermo Scientific) were thawed at room temperature, mixed vigorously for 5–10 s and centrifuged for 10 min at 10,000 × g at room temperature in a fixed angle rotor. The concentrations of Aβ1-40, Aβ1–42, pTau217 and GFAP were determined using the commercially available plasma Aβ1-40, Aβ1–42 (n = 230 each), pTau217 (n = 233) and GFAP (n = 228) Immunoreaction Cartridges on the fully automated Lumipulse G600II System [100]. Manufacturer quality control material was analyzed before and after testing each day and determined to be within the manufacturer’s specifications. Precision based on quality control data was < 5% CV for all Lumipulse assays tested. Internal quality control samples were included in all runs to assess inter-assay variability. EDTA plasma was introduced into the instrument using individual 2 mL screw cap micro tubes (Sarstedt, Germany). All assays were performed as single measurements following the kit instructions.

### Assessment of Physical Fitness

Physical fitness assessment included measurements of both aerobic (VO_2_max) and muscular capacity (inverse proxy of sarcopenia). However, due to time constraints in the later stages of the study, only muscular capacity was assessed (small physical fitness assessment), resulting in a larger sample size for this measure (N = 345). Consequently, data on both muscular and aerobic capacity were available for only a subsample of participants (N = 140; comprehensive physical fitness assessment).

Cardiorespiratory fitness was quantified using a symptom-limited cardiopulmonary exercise test (CPET) on a cycle ergometer (ergoselect 200, COSMED). The protocol commenced at 25 W, with incremental increases of 25 W every 3 minutes, flanked by warm-up and cool-down phases (0 W, 3 min each). Expired gases were continuously monitored via breath-by-breath analysis (Quark CPET), alongside heart rate (12-lead ECG), blood pressure, and subjective exertion (Borg Scale 6–20). Termination criteria included a VO₂ plateau, respiratory exchange ratio > 1.10, or volitional fatigue.

Muscular capacity was evaluated through a composite of maximal handgrip strength (SAEHAN dynamometer; average of three maximal voluntary contractions per hand), the timed-up-and-go-test (TUG), and appendicular skeletal muscle mass (ASMM) assessed via bioelectrical impedance analysis (BIA 101 BIVA PRO). A standardized z-score composite was derived from these measures, with TUG scores inverted (TUG x –1) to ensure consistent interpretation: higher scores reflected superior muscular function.

### Statistical Analyses

All statistical analyses were performed using R (version 4.3.2) within the RStudio environment (version 2023.9.1.494). Data visualization was conducted utilizing the ggplot2 package, as part of the tidyverse package [101], and the corrplot package [102].

Prior to conducting inferential analyses, the assumptions underlying linear models were rigorously assessed. Normality of residuals was examined via the Shapiro–Wilk test, complemented by visual inspection of histograms and Q–Q plots. Variables that were right-skewed were log-transformed prior to analysis and outlier removal. Extreme outliers were defined as data points exceeding ±3 times the interquartile range and were excluded from subsequent analyses. Linearity was evaluated by plotting residuals against fitted values, and log transformation was applied in cases of substantial skewness. Homoscedasticity was assessed using the Breusch–Pagan test, and the Durbin–Watson test was employed to verify the independence of residuals.

An alpha threshold of p < 0.05 was used for all hypothesis testing. Correction for multiple comparisons was implemented using the Benjamini–Hochberg False Discovery Rate (FDR) procedure to minimize the likelihood of Type I errors in the context of multiple inferential tests.

### Hypothesis 1: Association between Physical Fitness and Cognitive Performance

To investigate the relationship between physical fitness and cognitive functioning, linear regression models were constructed using the lm function from the stats package. Aerobic and muscular fitness were entered as independent variables in separate models, predicting cognitive performance outcomes across the domains assessed by the CERAD, PACC5, VLMT, and ROCFT. Covariates included age, sex, and years of education. Interaction terms of physical fitness indices and sex were initially incorporated to explore potential sex-specific effects. Non-significant interaction terms were removed to simplify model interpretation.

### Hypothesis 2: Physical Fitness and Resistance to Brain Pathology (Brain Maintenance)

To explore the contribution of physical fitness to neurobiological resilience, a series of regression models were estimated with markers of neuropathology as dependent variables. Predictors included aerobic fitness (VO₂max), muscular fitness (z-standardized composite score). Neuroimaging and biomarker outcomes included fractional BG and CSO PVS volume, whole-brain WMH volume, medial temporal lobe (MTL) tau load (DVR), plasma pTau217, and the Aβ₁₋₄₂/ Aβ₁₋₄₀ ratio. Age and sex were included as covariates in all models.

### Hypothesis 3: Physical Fitness as a Proxy for Cognitive Reserve

To examine the hypothesis that physical fitness serves as a proxy for cognitive reserve, a multistep analytical strategy was employed. First, associations between neuropathological markers (BG PVS, CSO PVS, WMH, MTL DVR, pTau217, and Aβ₁₋₄₂/ Aβ₁₋₄₀) and cognitive performance (PACC5, CERAD, VLMT, ROCFT) were assessed using linear regression (Figure 2A). CERAD and PACC5 were prioritized as global cognitive outcomes due to its established sensitivity to preclinical amyloid-related decline [84], while VLMT and ROCFT provided domain-specific measures of episodic memory, which is particularly susceptible to age-related deterioration [1,103]. Where a pathology marker exhibited a significant association with cognitive performance, interaction terms were added to assess whether physical fitness (VO₂max or muscular capacity) moderated this relationship, thus providing evidence for cognitive reserve (Figure 2B). A significant interaction was interpreted as robust evidence for cognitive reserve mechanisms.

**Fig 2.**
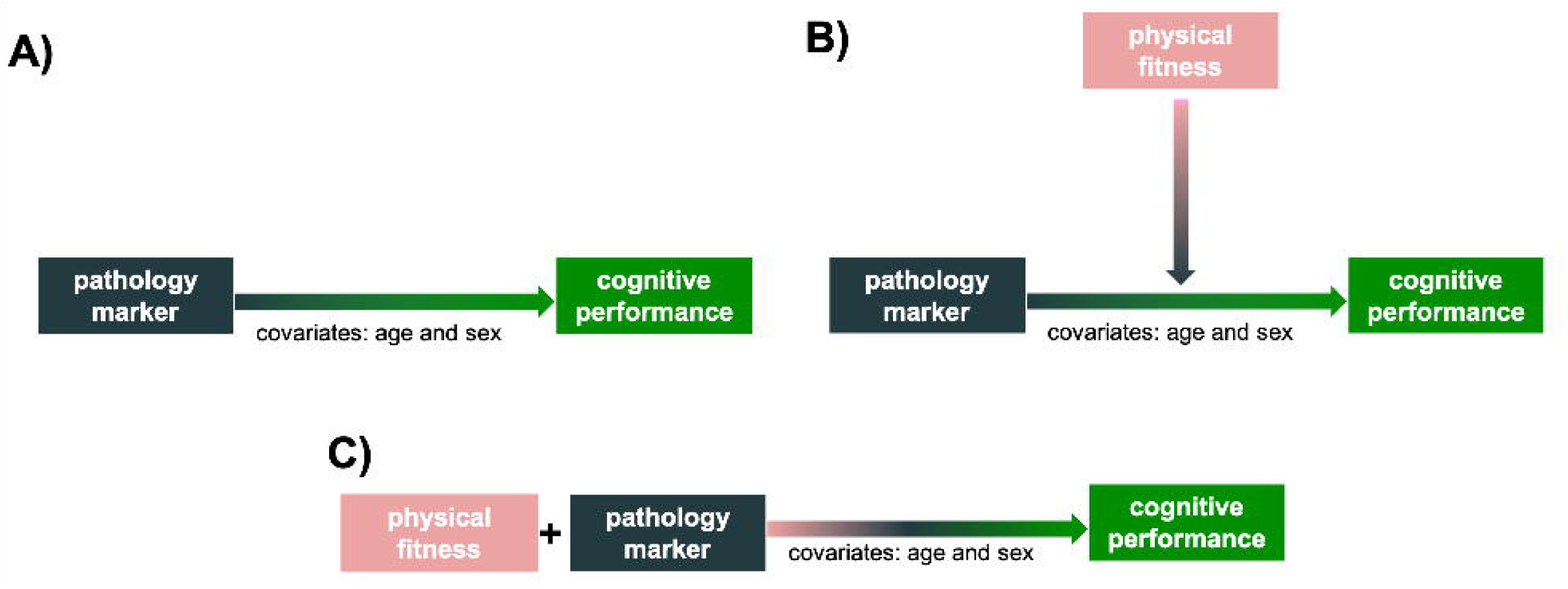
Cognitive reserve analysis. (A) Baseline-condition: influence of pathology markers on cognitive performance. (B) Moderation analysis: examining the interaction between physical fitness and pathology markers on cognitive outcomes. (C) Analysis for weak evidence of cognitive reserve: model including both physical fitness and pathology markers as predictors of cognitive performance. Cognitive performance measures: PACC5, CERAD, VLMT, ROCFT; pathology marker: BG PVS, CSO PVS, WMH, MTL DVR, ptau217, Aβ₁₋₄₂/Aβ₁₋₄₀; physical fitness markers: VO_2_max and muscular capacity; covariates: age (in years) and sex.

In cases where the interaction was not significant, but the fitness proxy independently predicted cognitive performance, a model comparison approach was applied. Specifically, models with and without the fitness variable were compared using ANOVA to determine whether its inclusion significantly improved model fit (Figure 2C), providing weaker yet supportive evidence for cognitive reserve [11]. All models controlled for age and sex. Education was excluded as a covariate due to its established role as a canonical proxy for cognitive reserve, which could confound interpretations [10,104].

### Hypothesis 4: Neurobiological Mechanisms Linking Physical Fitness and Cognitive Function

To investigate potential associations between hippocampal volume and peripheral neurotrophic biomarkers, Pearson correlation coefficients corrected for multiple comparisons using the Benjamini–Hochberg FDR procedure were computed. Specifically, correlations were examined between total, anterior, and posterior hippocampal volumes and serum concentrations of BDNF, VEGF, and Cathepsin B. Prior to analysis, the assumption of linearity was evaluated via visual inspection of scatterplots. Extreme outliers—defined as data points exceeding ±3 times the interquartile range—were excluded from subsequent analyses.

To further elucidate the potential neurobiological mechanisms underlying the observed associations between physical fitness and cognitive performance, mediation analyses were conducted (Figure 3). These analyses were implemented using the mediation package in R [105], with statistical inference based on a nonparametric bootstrapping approach (50,000 iterations) as recommended by [106,107]. Mediation models were estimated only when the proposed independent variables—namely, aerobic fitness (VO₂max) and muscular capacity—were significantly related to both the hypothesized mediators (left, right, and total anterior hippocampal volume [aHCV], posterior hippocampal volume [pHCV], whole hippocampal volume [wHCV], total grey matter volume [GMvol], and circulating neurotrophic factors) and the cognitive outcomes (PACC5, CERAD, VLMT, and ROCFT).

**Fig 3.**
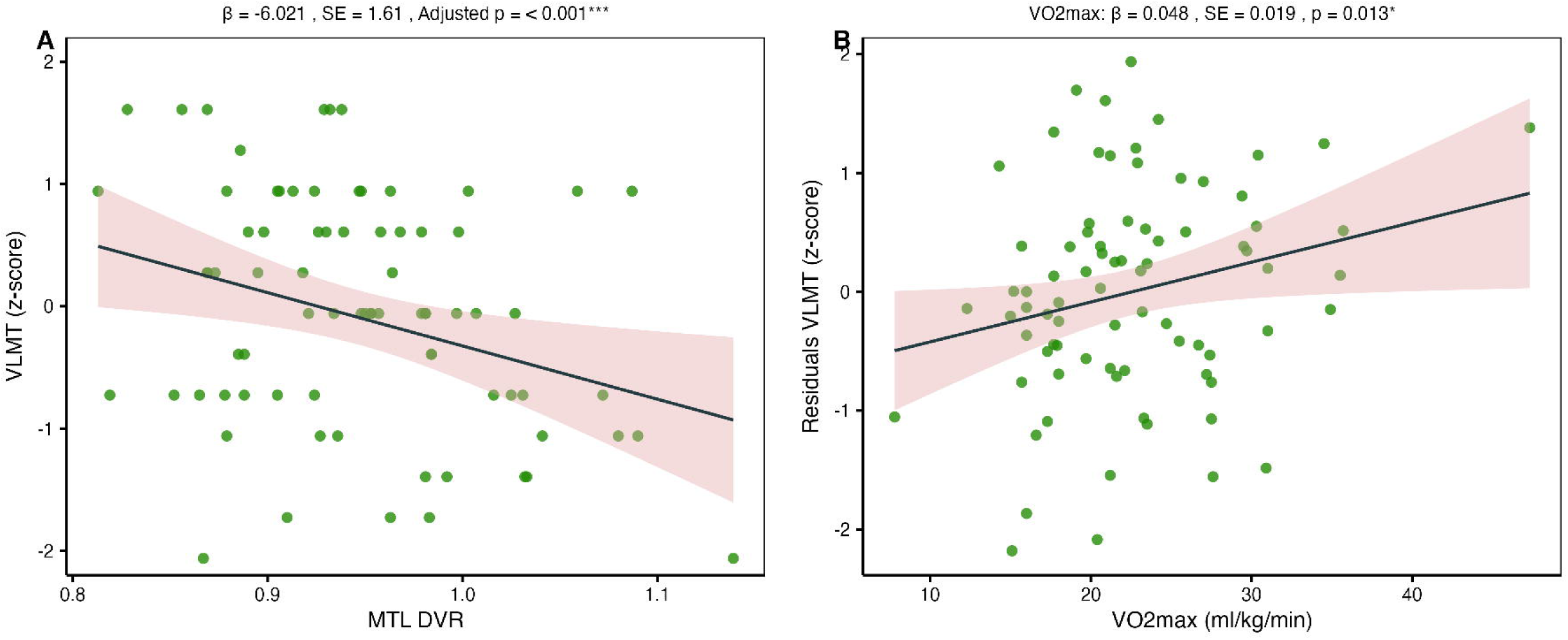
Mediation analysis examining the association between physical fitness and cognitive performance. (A) Total effect (c): the overall effect of physical fitness markers (VO_2_max and muscular capacity, separately) on cognitive performance measures (PACC5, CERAD, VLMT, ROCFT). (B) Mediation decomposition: the indirect effect (average causal mediation effect, a*b) represents the portion of the effect transmitted through the mediator, while the direct effect (c’) represents the remaining effect of physical fitness on cognition not explained by the mediator. The total effect is the sum of the direct and indirect effects, expressed as c=c’+(a∗b).

All mediation models were adjusted for age, sex, and years of education. TIV was not included as a covariate, given that all volumetric brain measures had been pre-adjusted for TIV during preprocessing.

## Results

A total of 345 cognitively normal older participants (177 females) were included in the study (age: M = 72.77 years, SD = 7.95; years of education: M = 14.93 years, SD = 2.38). The Supplemental Material provides additional uncorrected raw baseline values for the physical fitness outcomes (Table S1), cognitive outcomes and pathology markers (Table S2), as well as the structural volumetric parameters (Table S3). Muscular capacity and VO₂max were significantly positively correlated (ρ = 0.326, p < .001; Fig. 4), indicating that individuals with higher muscular capacity also tended to exhibit higher aerobic capacity.

**Fig 4.**
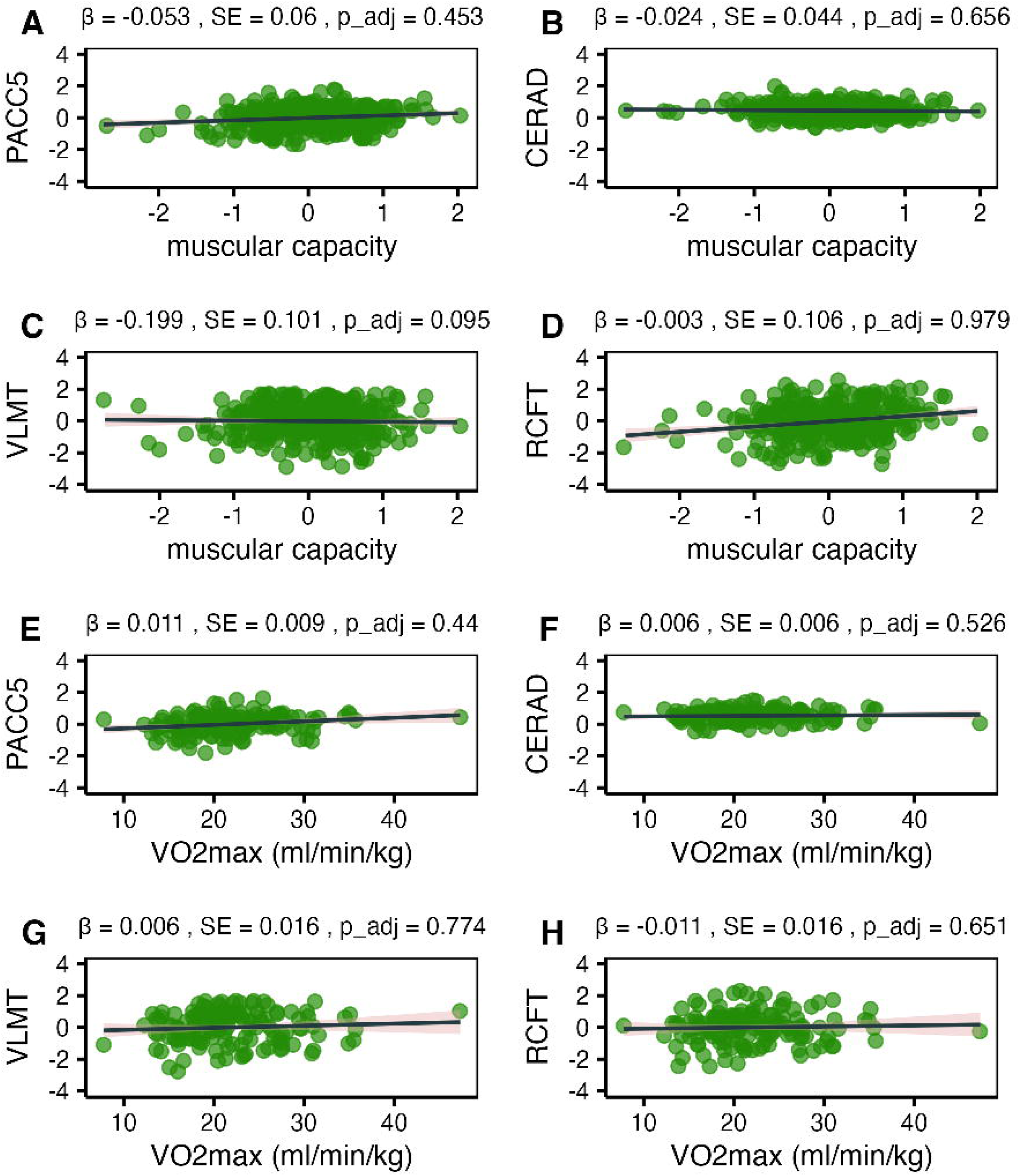
Scatter plots of bivariate associations between physical fitness outcomes. Panel A shows VO_2_max (aerobic capacity, in ml/min/kg) and muscular capacity (z-composite: TUG, maximal handgrip strength and ASMM). Panels B-D show the muscular capacity outcomes separately. A linear trend line is shown with a 95% confidence interval (light pink area) to illustrate the relationship between the variables. ρ, Spearman’s rank correlation coefficient; Adjusted p: FDR-corrected (False Discovery Rate) p-value (Benjamini-Hochberg procedure).

Within the muscular capacity composite, associations among its constituent measures, TUG performance, maximal handgrip strength, and ASMM, were evaluated (all z-standardized). Outliers were excluded from TUG (n = 4) and ASMM (n = 1). Higher TUG performance was positively correlated with maximal handgrip strength (ρ = 0.427, p < .001; Fig. 4). In contrast, TUG performance was not significantly correlated with ASMM, nor was maximal handgrip strength with ASMM (p > .05; Fig. 4).

### Hypothesis 1: Association between Physical Fitness and Cognitive Performance

To account for potential sex differences, separate models for each cognitive outcome initially included a physical fitness by sex interaction term. No significant interactions were observed for either VO₂max or muscular capacity (p > .05 for both).

Next, the association between physical fitness and cognitive performance was tested (Fig. 5). Separate regression models were specified for each cognitive outcome. Muscular capacity was not significantly associated with global cognitive performance (PACC5: β = −0.053, SE = 0.06, p_adjusted_ = .453; CERAD: β = −0.024, SE = 0.044, p_adjusted_ = .656) or episodic memory performance (VLMT: β = −0.199, SE = 0.101, p_adjusted_ = .088; ROCFT: β = −0.003, SE = 0.106, p_adjusted_ = .979). Similarly, VO₂max did not significantly predict global cognition (PACC5: β = 0.011, SE = 0.009, p_adjusted_ = .440; CERAD: β = 0.006, SE = 0.096, p_adjusted_ = .526) or episodic memory (VLMT: β = 0.006, SE = 0.016, p_adjusted_ = .774; ROCFT: β = −0.011, SE = 0.016, p_adjusted_ = .651).

**Fig 5.**
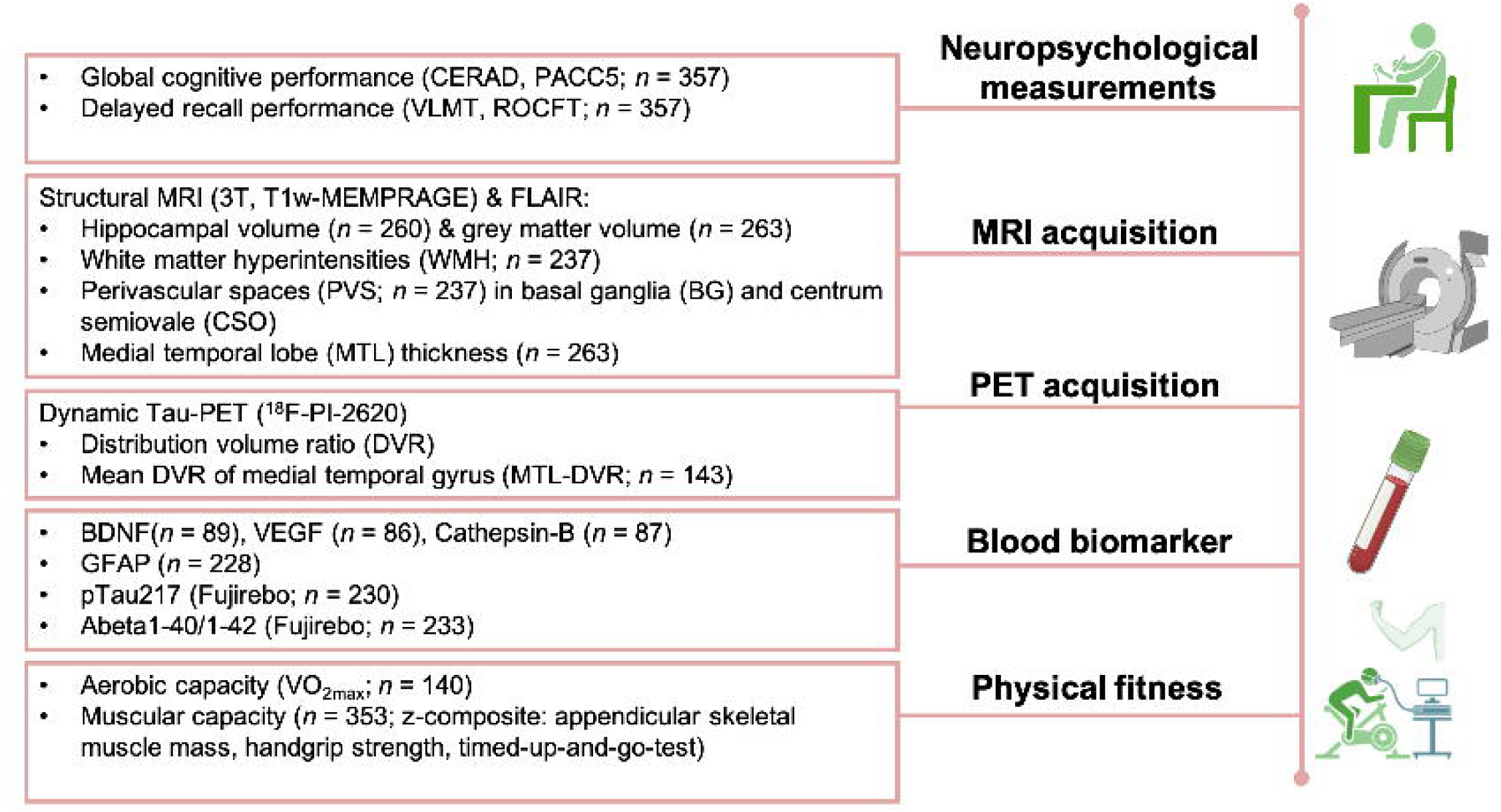
Linear regression plots of physical fitness and cognitive performance (raw z-score values). Physical fitness: muscular capacity (z-composite: TUG, maximal handgrip strength and ASMM; panels A - D) and VO_2_max (aerobic capacity, in ml/min/kg; panels E - H); cognitive performance: CERAD, Consortium to establish a Registry for Alzheimer’s Disease Plus version (z-composite); PACC5, preclinical Alzheimer’s composite (z-composite); VLMT, German adaptation of Rey’s Auditory Verbal Learning Test (delayed verbal memory performance); ROCFT, Rey-Osterrieth Complex Figure Test (visuospatial delayed memory performance). Estimated regression line is shown with a 95% confidence interval (light pink area). Β, beta coefficient (slope); SE: standard error; Adjusted p: FDR-corrected (False Discovery Rate) p-value (Benjamini-Hochberg procedure); All models included age, sex and years of education as covariates.

Hypothesis 1 was not supported, as no significant associations were observed between physical fitness and cognitive performance.

### Hypothesis 2: Physical Fitness and Resistance to Brain Pathology (Brain Maintenance)

For both physical fitness samples (muscular capacity: N = 345; aerobic capacity subsample: N = 140), outliers were excluded from pTau217 (muscular capacity: n = 4; aerobic capacity: n = 1) and Aβ₁₋₄₂/ Aβ₁₋₄₀ (muscular capacity: n = 1). To achieve normal distributions, Aβ₁₋₄₂/ Aβ₁₋₄₀, GFAP, and pTau217 values were log-transformed before outlier removal. Separate analyses were conducted within each physical fitness sample to predict markers of neuropathology, including BG and CSO PVS, WMH, and, in the muscular capacity sample, MTL tau, pTau217, Aβ₁₋₄₂/ Aβ₁₋₄₀, and GFAP.

A significant negative association was observed between muscular capacity and BG PVS (β = −0.384, SE = 0.136, p_adjusted_ = .010; Fig. 6A), such that higher muscular capacity was linked to lower BG PVS volumes. For VO₂max, a negative association with BG PVS was also observed (p = .049), but this did not survive FDR correction (β = −0.029, SE = 0.015, p_adjusted_ = .089; Fig. 6A). No significant associations were found between either physical fitness measure and other pathology markers (all p_adjusted_ > .05). Detailed associations between individual muscular capacity components and pathology markers are provided in the Supplemental Material (Fig. S1).

**Fig 6.**
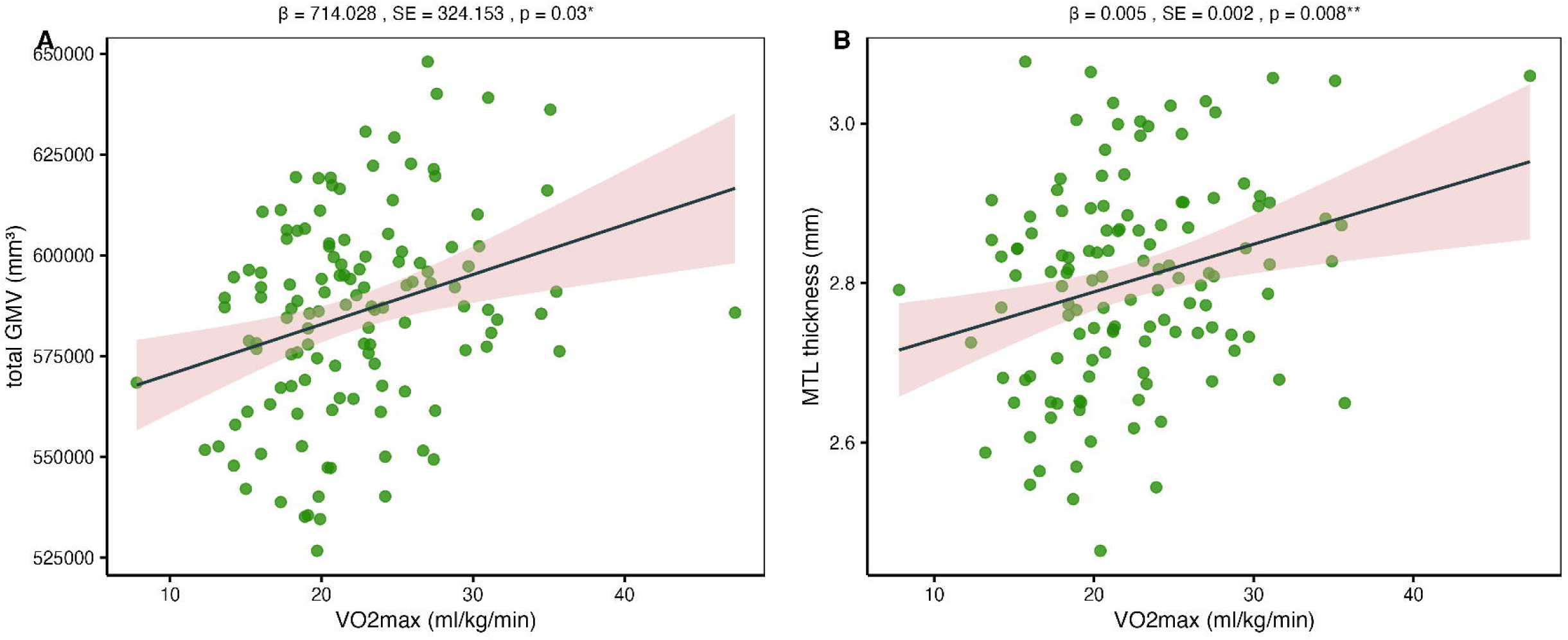
Linear regression plots illustrating the association between physical fitness and pathology markers. Left: effect of muscular capacity (z-standardized composite of TUG, ASMM, and maximal handgrip strength) on BG PVS volumes (basal ganglia perivascular spaces, normalized for BG volume). Right: effect of VO₂max (aerobic capacity) on BG PVS volumes. The estimated regression line is displayed with its 95% confidence interval (light pink shaded area). β: regression slope (beta coefficient); SE: standard error; Adjusted p: FDR-corrected p-value (Benjamini–Hochberg procedure). * Indicates Adjusted p < .05. All models were adjusted for age and sex.

In summary, partial support for Hypothesis 2 [brain maintenance] was observed: higher muscular capacity (and aerobic capacity at trend-level) predicted reduced BG PVS volumes, whereas no significant associations were found for the other pathology markers assessed.

### Hypothesis 3: Physical Fitness as a Proxy for Cognitive Reserve

First, associations between pathology markers and cognitive performance were evaluated. In the muscular capacity subgroup, a positive association was observed between Aβ₁₋₄₂/ Aβ₁₋₄₀ and PACC5 scores (β = 0.884, SE = 0.294, p_adjusted_ = .006), such that less pathological plasma Aβ levels related to better cognitive performance. No significant relationships were detected between any other pathology markers and cognitive outcomes (p_adjusted_ > .05). Muscular capacity was subsequently tested as a potential cognitive reserve proxy. No significant interaction was observed between muscular capacity and Aβ₁₋₄₂/Aβ₁₋₄₀ (β = 5.95, SE = 5.58, p_adjusted_ = .287), indicating that muscular capacity did not moderate the association between Aβ₁₋₄₂/ Aβ₁₋₄₀ levels and global cognitive performance. In a subsequent model including both muscular capacity and Aβ₁₋₄₂/ Aβ₁₋₄₀ as predictors, muscular capacity did not predict PACC5 performance (β = 0.031, SE = 0.087, p = .717), providing no evidence for even a weak cognitive reserve effect.

Within the VO₂max subgroup, higher MTL DVR values were significantly associated with poorer delayed verbal memory on the VLMT (β = −6.02, SE = 1.61, p_adjusted_ < .001), indicating that elevated tau pathology is linked to reduced delayed memory performance (Fig. 7A). No significant associations were detected between pathology markers and PACC5, CERAD, or ROCFT scores in this subgroup (all p_adjusted_ > .05). VO₂max was then evaluated as a potential cognitive reserve proxy. No significant interaction was observed between VO₂max and MTL DVR (β = −0.05, SE = 0.20, p_adjusted_ = .780), indicating that VO₂max does not moderate the relationship between tau pathology and delayed verbal memory.

**Fig 7.**
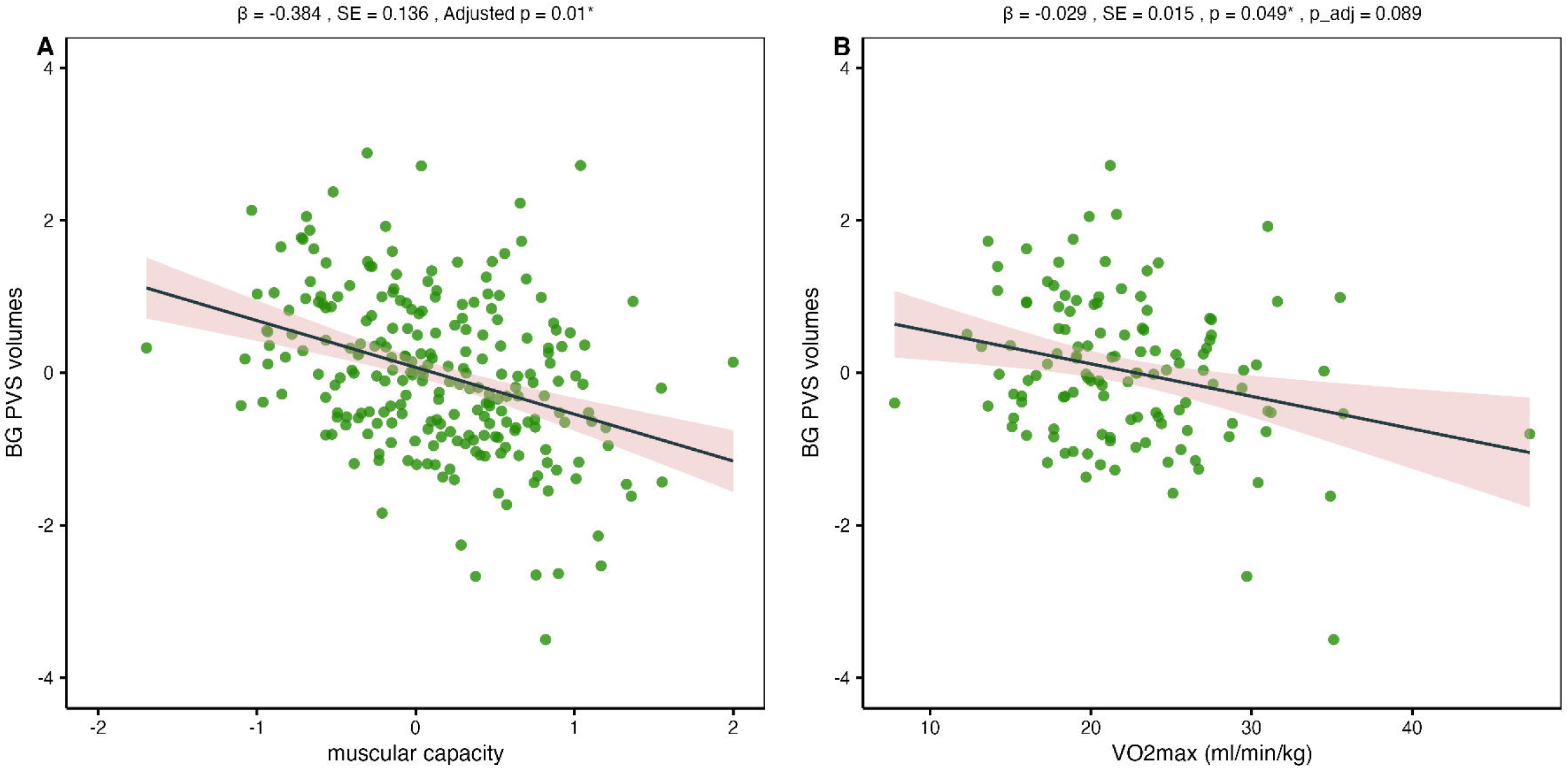
**A:** Association between medial temporal lobe Tau burden (MTL DVR; predictor) and delayed verbal memory performance (VLMT z-score; dependent variable), adjusted for age and sex. **B:** Association between VO₂max (aerobic capacity as predictor, ml/min/kg) and delayed verbal memory performance (VLMT, dependent variable) after removal of variance attributable to age, sex, and MTL DVR Tau. VLMT residuals were regressed on VO₂max to illustrate weak evidence for cognitive reserve. The estimated regression line is displayed with its 95% confidence interval (light pink shaded area). β: regression slope (beta coefficient); SE: standard error; Adjusted p: FDR-corrected p-value (Benjamini– Hochberg procedure). * Indicates Adjusted p < .05.

However, in a model including both VO₂max and MTL DVR as predictors, VO₂max was positively associated with VLMT performance (β = 0.048, SE = 0.019, p = .013; Fig. 7B). Model comparison revealed a significantly improved fit for the combined model relative to the model excluding VO₂max (F(1,74) = 6.48, p = .012), suggesting that VO₂max explains additional variance in delayed verbal memory beyond that accounted for by MTL tau pathology alone.

Taken together, Hypothesis 3 is partially supported. Weak evidence for aerobic fitness (VO₂max) as a proxy for cognitive reserve was limited: VO₂max independently predicted better VLMT performance but did not moderate the association between pathology and cognition. No evidence was found supporting muscular capacity as a proxy for cognitive reserve.

### Hypothesis 4: Neurobiological Mechanisms Linking Physical Fitness and Cognitive Function

As no significant associations were observed between either aerobic or muscular capacity and any cognitive outcomes (p > .05; see Hypothesis 1, Fig. 5), no mediation analyses were conducted for total effects (path c). Path a, representing the association between physical fitness and potential mediators was examined separately for each physical fitness modality. Potential mediators included bilateral, left, and right aHCV, pHCV, wHCV, total GMV, MTL thickness, and neuroplasticity markers BDNF, VEGF, and Cathepsin-B.

In the VO₂max sample, positive associations were observed between VO₂max and total GMV (β = 714.03, SE = 324.15, p = .03; Fig. 8A) and MTL thickness (β = 0.005, SE = 0.002, p = .008; Fig. 8B), whereas no other significant relationships with potential mediators were detected. In the muscular capacity sample, a negative association emerged between muscular capacity and right aHCV (β = −80.32, SE = 33.29, p_adjusted_ = .033), with no significant associations observed for other hippocampal volumes, MTL thickness, or plasticity-related markers.

**Fig 8.**
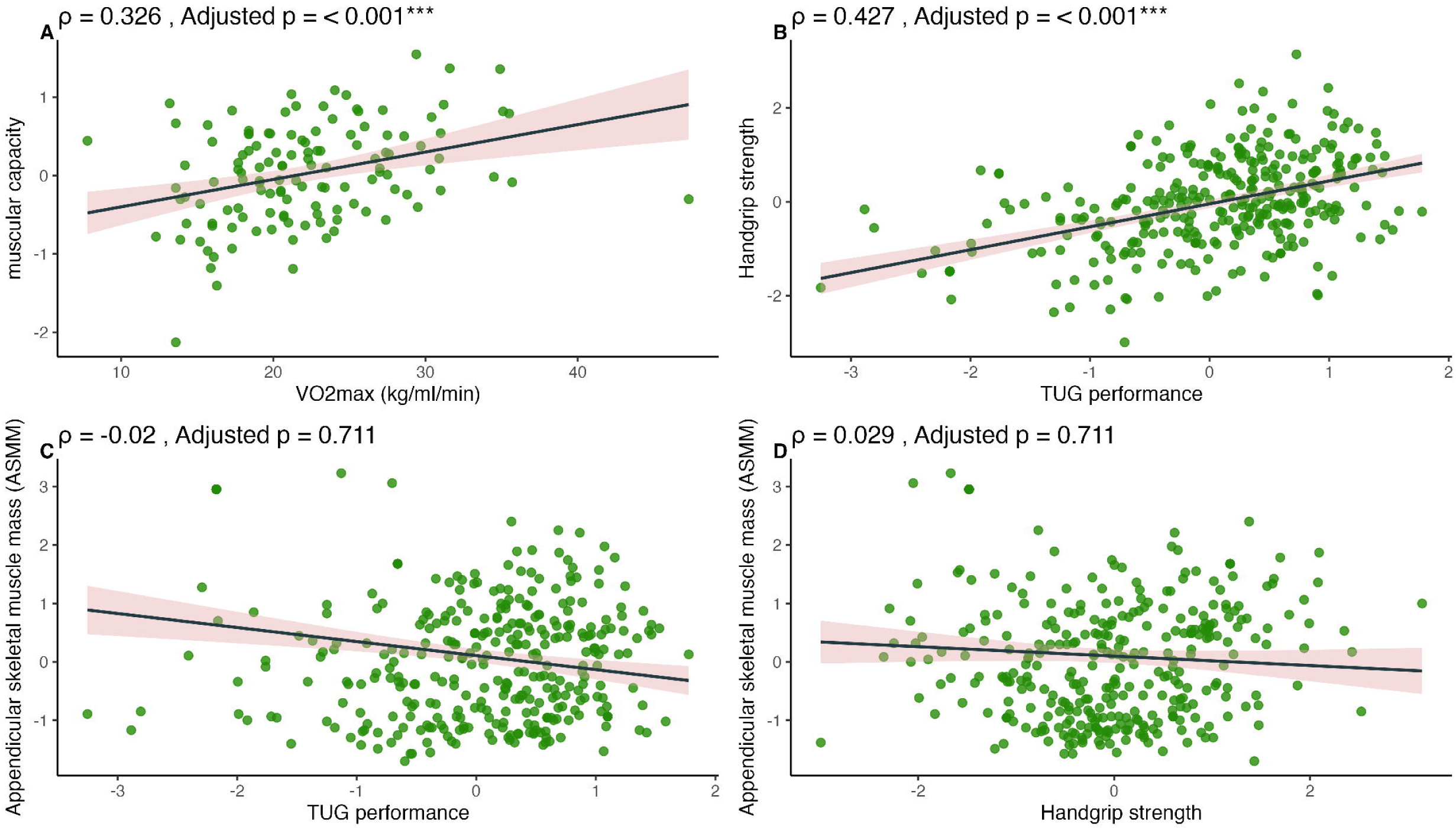
Potential mediators – multiple linear regression plots of path a. A: Effect of VO_2_max (aerobic capacity as predictor, ml/min/kg) on total gray matter volume (GMV, mm^3^) and B: on Medial temporal lobe (MTL) thickness (in mm). The estimated regression line is displayed with its 95% confidence interval (light pink shaded area). β: regression slope (beta coefficient); SE: standard error; Adjusted p: FDR-corrected p-value (Benjamini–Hochberg procedure). * indicates Adjusted p < .05. All models were adjusted for age and sex (raw values are plotted).

## Discussion

This study examined the role of physical fitness, specifically aerobic capacity (VO₂max) and muscular capacity (used inversely as a proxy for sarcopenia), with respect to cognitive performance, brain outcomes, and markers of pathology among cognitively normal older adults. There was no association between either physical fitness modality and cognitive performance, and, no mediating effect of hippocampal volume, MTL thickness, total GMV, or plasticity-associated factors. Nevertheless, a higher aerobic capacity was related to greater MTL thickness and total GMV, suggesting a beneficial or potentially protective effect on brain structure. There was further evidence that muscular capacity contributed to brain maintenance, as indicated by a negative association with BG PVS volumes, suggesting potential resistance to brain pathology. Furthermore, the study provided weak evidence that aerobic fitness might contribute to cognitive reserve against the detrimental effect of MTL tau pathology on verbal memory performance. Specifically, at similar levels of tau pathology, individuals with higher aerobic capacity demonstrated better verbal memory performance. These findings suggest that physical fitness may contribute to resilience to age-related cognitive decline by enhancing cognitive reserve and resistance to neuropathological processes.

To the best of our knowledge, this is the first study showing that higher muscular capacity is linked to reduced PVS volumes in the basal ganglia. We also observed a similar, albeit weaker, association for aerobic fitness in a smaller subsample, which did not remain after FDR correction. PVS plays an essential role in the clearance of metabolic and neurotoxic waste, including Aβ and tau, and its enlargement may reflect impaired glymphatic function [108–112]. In aged mice, voluntary wheel running enhances glymphatic efficiency by maintaining perivascular AQP4 polarization and promoting Aβ clearance [113,114]. Although no human studies have directly examined the effects of aerobic exercise on PVS, it is plausible that aerobic activity mitigates age- and cardiovascular-related alterations through improved cerebrovascular health, enhanced glymphatic clearance, and reduced inflammation [115–117]. Given that enlargement of BG PVS has been associated with cardiovascular pathology [113], the lower BG PVS volumes observed in individuals with greater muscular capacity may reflect superior vascular integrity, thereby supporting the concept of brain maintenance. Moreover, in a follow-up analysis on the individual measures of the muscular capacity composite, only poorer walking performance (TUG) was linked to increased BG PVS volumes. TUG performance, a marker of sarcopenia, has been associated with cognitive decline [73,74], suggesting that muscular fitness and other exercise modalities warrant further investigation as potential contributors to brain maintenance. Future studies should further investigate potential mechanisms, such as improved neurotoxin clearance glymphatic efficiency, and preserved cerebrovascular integrity.

However, with respect to plasma markers of Alzheimer’s pathology (p-tau217, Aβ₁₋₄₂/ Aβ₁₋₄₀) and GFAP, we found no significant associations with our objective measures of aerobic and muscular fitness. Similarly, a recent analysis in participants from the population-based Rotterdam Study also reported no association between accelerometer-measured physical activity and the same plasma AD biomarkers or PET measures of cortical Aβ 7 years later [118]. In a multicenter study in older adults with memory complaints, self-reported moderate-to-vigorous physical activity was likewise not associated with baseline p-tau181 levels but predicted slower changes in p-tau181 concentrations over 3 years [34]. The association between physical activity and GFAP levels - a biomarker of astrocyte activation and an early marker of Aβ pathology [119] - remains insufficiently investigated. In rats, exercise has been shown to induce increases in GFAP-positive astrocytes within the hippocampal CA1 region [120]. In humans, however, findings are less consistent: some studies reported cross-sectional associations between higher levels of physical activity and lower GFAP concentrations, indicative of reduced astrocytic inflammation and degeneration in older adults [121], whereas others [122], including our study, did not observe such association. Overall, findings have been mixed [31,122], with some studies reporting no associations (see also [123]), whereas other show a beneficial link between fitness measures and cerebrospinal fluid (CSF) AD biomarkers [124–127] in different samples. These discrepancies may be partly explained by differences in physical activity and AD biomarker assessment, sample characteristics (e.g., exclusion criteria, cognitive status), and adjustment for confounding factors.

Additionally, there was weak evidence suggesting that greater aerobic capacity may confer or contribute to greater cognitive reserve, particularly against tau-related memory deficits. Tau pathology, a defining feature of AD, is also common in cognitively intact older adults, with deposition initiating in the MTL an area critical for episodic memory [128–132]. According to Braak staging, tau aggregation begins in the transentorhinal cortex and advances through the entorhinal cortex, hippocampus, perirhinal cortex, lateral temporal regions, and subsequently the frontal and parietal cortices [128,133]. In our study, other pathological markers, including global WMH, PVS, and AD-related blood biomarkers (Aβ₁₋₄₂/ Aβ₁₋₄₀, p-tau217, and GFAP), were not associated with cognitive outcomes. These markers, except PVS, represent more global indicators of pathology, whereas MTL tau deposition has been consistently linked to episodic memory impairment and regional atrophy in cognitively normal aging [131,134–138]. Hence, the observed association between MTL tau and memory performance confirms prior literature. Furthermore, evidence for a moderating role of physical activity on tau-cognition associations also comes from a recent study in non-demented older adults, where the negative relationship between CSF tau measures (p-tau181 and total tau) and executive function was attenuated among more physically active participants [139]. Overall, evidence supporting the role of physical fitness as a moderating factor in the relationship between AD pathology and cognition highlights the potential of exercise-based interventions to mitigate tau-related vulnerability and enhance cognitive reserve mechanisms.

Although the causal mechanisms underlying physical activity’s cognitive benefits remain under debate, converging evidence indicates that exercise enhances brain function through molecular, cellular, structural, and systemic processes [24,140]. Animal studies consistently demonstrate that voluntary running promotes hippocampal neurogenesis, long-term potentiation, BDNF upregulation, and hippocampus-dependent memory, even in AD models [52,54,141–143]. Exercise also stimulates the secretion of peripheral myokines, such as irisin and Cathepsin B, which can cross the blood–brain barrier and induce BDNF synthesis, thereby fostering synaptic plasticity and cognitive maintenance [55,56,59–61,144,145]. Additionally, anaerobic exercise– induced lactate may cross the blood–brain barrier and activate HCAR1, leading to VEGF-mediated angiogenesis and enhanced neurovascular support for cognition [68,69]. However, translating these mechanisms to humans remains challenging. While some cross-sectional studies report hippocampal volume or plasticity-related factors as mediators between aerobic fitness and cognition [50,146,147], others - including the present study - did not confirm these relationships [24,47,49,148].

In this cohort, physical fitness was not associated with cognitive performance, hippocampal volume, or plasticity-related factors, and no mediating effects were detected. This may reflect sample homogeneity, limiting variance in cognitive and brain measures, or suggest that hippocampal volume alone does not capture distributed exercise-induced neural adaptations. Improvements in vascular function, network connectivity, or systemic signaling may play more prominent roles in sustaining cognition [14,57]. Nevertheless, VO₂max was positively related to MTL thickness and total GMV, suggesting subtle neuroprotective effects that could preserve cognition in normal aging. Future longitudinal studies combining multimodal imaging, biomarker assessment, and diverse physical exercise interventions are needed to elucidate the pathways through which physical activity supports brain health.

The use of objective measures of physical fitness (VO₂max and muscular capacity) constitutes a methodological advantage over studies relying on subjective or indirect assessments. However, the cross-sectional design limits causal inference and constrains the evaluation of brain maintenance, which is best examined through longitudinal approaches [10,11]. Nonetheless, such cross-sectional data provide valuable insights into associations between fitness and AD-related pathology markers, thereby informing preventive and clinical intervention strategies. Thus, further longitudinal and interventional studies are needed to determine whether, and which modalities of, physical exercise training can directly reduce BG PVS burden and enhance cognitive outcomes.

This study investigated the role of different physical fitness modalities in normal cognitive aging, with a focus on their potential contribution to resilience. However, several limitations should be noted. The sample consisted of cognitively normal, highly educated, community-dwelling Caucasian German individuals, introducing potential selection bias and limiting generalizability. The relatively small sample size with measures of aerobic capacity reduced statistical power, and the cross-sectional design precluded causal inference. Physical fitness was assessed at a single time point, which may not capture temporal variability. Other domains of physical fitness, such as balance, flexibility, and functional training, were not evaluated, though they may also influence cognitive health.

## Conclusion

Higher muscular capacity was associated with reduced basal ganglia PVS volumes, supporting structural brain maintenance and healthy aging. Aerobic fitness (VO₂max) showed a weak link to cognitive reserve, particularly in the context of MTL tau pathology and delayed verbal memory, despite no direct association with global cognitive performance. Potential mediators—including hippocampal volume and BDNF—did not explain these relationships. VO₂max was positively associated with total GMV and MTL thickness, highlighting a potential neuroprotective effect. These findings suggest that physical fitness may enhance resilience to AD-related neuropathology, promote cognitive health, and contribute to the extension of healthspan. Longitudinal, mechanistic studies are required to clarify how physical exercise supports cognitive health.

## Declarations

### Ethics approval and consent to participate

The study design and protocol were approved by the local Ethics Committee of the Medical Faculty, Otto-von-Guericke University Magdeburg (200/19) and were carried out in accordance with the code of ethics of the World Medical Association (Declaration of Helsinki, 1967). Written informed consent was obtained from all participants.

### Consent for publication

Not applicable.

### Availability of data and materials

All data supporting the findings of this study are reported within the article and its Supplementary Information. The data were derived from the SFB 1436 study (Z03 cohort). Due to ethical and data protection regulations governing this cohort, the underlying individual-level data are not publicly available. Access to the data may be possible upon reasonable request and subject to approval by the SFB 1436 data governance committee.

## Data Availability

All data produced in the present study are available upon reasonable request to the authors

## Acknowledgements

We would like to express our gratitude to our test subjects, who, with great curiosity, patience and commitment, made our research possible in the first place. We further thank Kathrin Baldauf and Peter Schulze for their assistance and support regarding the MRI scanning. We also would like to thank the medical-technical assistants (MTAs), in particular Ulrike Pankratz, Vivica Sommerfeld and Iris Mann, as well as our student assistants for their valuable contributions to this study.

## References

1. Brito DVC, Esteves F, Rajado AT, Silva N, Andrade R, Apolónio J, et al. Assessing cognitive decline in the aging brain: lessons from rodent and human studies. npj Aging [Internet]. 2023;9:23. 10.1038/s41514-023-00120-6

2. Franceschi C, Garagnani P, Morsiani C, Conte M, Santoro A, Grignolio A, et al. The Continuum of Aging and Age-Related Diseases: Common Mechanisms but Different Rates. Front Med (Lausanne) [Internet]. 2018;5. 10.3389/fmed.2018.00061

3. Li Z, Zhang Z, Ren Y, Wang Y, Fang J, Yue H, et al. Aging and age-related diseases: from mechanisms to therapeutic strategies. Biogerontology [Internet]. 2021;22:165–87. 10.1007/s10522-021-09910-5

4. Lindenberger U. Human cognitive aging: Corriger la fortune? Science (1979) [Internet]. 2014;346:572–8. 10.1126/science.1254403

5. Raz N, Rodrigue KM. Differential aging of the brain: Patterns, cognitive correlates and modifiers. Neurosci Biobehav Rev [Internet]. 2006;30:730–48. 10.1016/j.neubiorev.2006.07.001

6. Sander MC, Fandakova Y, Werkle-Bergner M. Effects of age differences in memory formation on neural mechanisms of consolidation and retrieval. Semin Cell Dev Biol [Internet]. 2021;116:135–45. 10.1016/j.semcdb.2021.02.005

7. Salthouse TA. Decomposing age correlations on neuropsychological and cognitive variables. Journal of the International Neuropsychological Society [Internet]. 2009;15:650–61. 10.1017/S1355617709990385

8. Garo-Pascual M, Gaser C, Zhang L, Tohka J, Medina M, Strange BA. Brain structure and phenotypic profile of superagers compared with age-matched older adults: a longitudinal analysis from the Vallecas Project. Lancet Healthy Longev [Internet]. 2023;4:e374–85. 10.1016/S2666-7568(23)00079-X

9. Nyberg L, Pudas S. Successful Memory Aging. Annu Rev Psychol [Internet]. 2019;70:219–43. 10.1146/annurev-psych-010418-103052

10. Stern Y, Arenaza-Urquijo EM, Bartrés-Faz D, Belleville S, Cantilon M, Chetelat G, et al. Whitepaper: Defining and investigating cognitive reserve, brain reserve, and brain maintenance. Alzheimer’s & Dementia [Internet]. 2020;16:1305–11. 10.1016/j.jalz.2018.07.219

11. Stern Y, Albert M, Barnes CA, Cabeza R, Pascual-Leone A, Rapp PR. A framework for concepts of reserve and resilience in aging. Neurobiol Aging [Internet]. 2023;124:100–3. 10.1016/j.neurobiolaging.2022.10.015

12. Aghjayan SL, Bournias T, Kang C, Zhou X, Stillman CM, Donofry SD, et al. Aerobic exercise improves episodic memory in late adulthood: a systematic review and meta-analysis. Communications Medicine [Internet]. 2022;2:15. 10.1038/s43856-022-00079-7

13. Colcombe S, Kramer AF. Fitness Effects on the Cognitive Function of Older Adults. Psychol Sci [Internet]. 2003;14:125–30. 10.1111/1467-9280.t01-1-01430

14. Erickson KI, Donofry SD, Sewell KR, Brown BM, Stillman CM. Cognitive Aging and the Promise of Physical Activity. Annu Rev Clin Psychol [Internet]. 2022;18:417–42. 10.1146/annurev-clinpsy-072720-014213

15. Cabeza R, Albert M, Belleville S, Craik FIM, Duarte A, Grady CL, et al. Maintenance, reserve and compensation: the cognitive neuroscience of healthy ageing. Nat Rev Neurosci. 2018;19:701–10. 10.1038/s41583-018-0068-2

16. Jack CR, Wiste HJ, Weigand SD, Therneau TM, Knopman DS, Lowe V, et al. Age-specific and sex-specific prevalence of cerebral β-amyloidosis, tauopathy, and neurodegeneration in cognitively unimpaired individuals aged 50–95 years: a cross-sectional study. Lancet Neurol [Internet]. 2017;16:435–44. 10.1016/S1474-4422(17)30077-7

17. Boyle PA, Yu L, Wilson RS, Schneider JA, Bennett DA. Relation of neuropathology with cognitive decline among older persons without dementia. Front Aging Neurosci [Internet]. 2013;5. 10.3389/fnagi.2013.00050

18. De Silva TM, Faraci FM. Contributions of Aging to Cerebral Small Vessel Disease. Annu Rev Physiol [Internet]. 2020;82:275–95. 10.1146/annurev-physiol-021119-034338

19. Wardlaw JM, Benveniste H, Nedergaard M, Zlokovic B V., Mestre H, Lee H, et al. Perivascular spaces in the brain: anatomy, physiology and pathology. Nat Rev Neurol [Internet]. 2020;16:137–53. 10.1038/s41582-020-0312-z

20. Sanchez JS, Becker JA, Jacobs HIL, Hanseeuw BJ, Jiang S, Schultz AP, et al. The cortical origin and initial spread of medial temporal tauopathy in Alzheimer’s disease assessed with positron emission tomography. Sci Transl Med. 2021;13. 10.1126/scitranslmed.abc0655

21. Chauveau L, Kuhn E, Palix C, Felisatti F, Ourry V, de La Sayette V, et al. Medial Temporal Lobe Subregional Atrophy in Aging and Alzheimer’s Disease: A Longitudinal Study. Front Aging Neurosci. 2021;13. 10.3389/fnagi.2021.750154

22. Berron D, Vogel JW, Insel PS, Pereira JB, Xie L, Wisse LEM, et al. Early stages of tau pathology and its associations with functional connectivity, atrophy and memory. Brain. 2021;144:2771–83. 10.1093/brain/awab114

23. Erickson KI, Hillman C, Stillman CM, Ballard RM, Bloodgood B, Conroy DE, et al. Physical Activity, Cognition, and Brain Outcomes: A Review of the 2018 Physical Activity Guidelines. Med Sci Sports Exerc [Internet]. 2019;51:1242–51. 10.1249/MSS.0000000000001936

24. Stillman CM, Cohen J, Lehman ME, Erickson KI. Mediators of Physical Activity on Neurocognitive Function: A Review at Multiple Levels of Analysis. Front Hum Neurosci [Internet]. 2016;10. 10.3389/fnhum.2016.00626

25. Gow AJ, Bastin ME, Muñoz Maniega S, Valdés Hernández MC, Morris Z, Murray C, et al. Neuroprotective lifestyles and the aging brain. Neurology. 2012;79:1802–8. 10.1212/WNL.0b013e3182703fd2

26. Banerjee G, Kim HJ, Fox Z, Jäger HR, Wilson D, Charidimou A, et al. MRI-visible perivascular space location is associated with Alzheimer’s disease independently of amyloid burden. Brain [Internet]. 2017;140:1107–16. 10.1093/brain/awx003

27. Menze I, Bernal J, Kaya P, Aki Ç, Pfister M, Geisendörfer J, et al. Perivascular space enlargement accelerates in ageing and Alzheimer’s disease pathology: evidence from a three-year longitudinal multicentre study. Alzheimers Res Ther [Internet]. 2024;16:242. 10.1186/s13195-024-01603-8

28. Sepehrband F, Barisano G, Sheikh-Bahaei N, Choupan J, Cabeen RP, Lynch KM, et al. Volumetric distribution of perivascular space in relation to mild cognitive impairment. Neurobiol Aging [Internet]. 2021;99:28–43. 10.1016/j.neurobiolaging.2020.12.010

29. Marino FR, Deal JA, Dougherty RJ, Bilgel M, Tian Q, An Y, et al. Differences in Daily Physical Activity by Alzheimer’s Risk Markers Among Older Adults. J Gerontol A Biol Sci Med Sci. 2024;79. 10.1093/gerona/glae119

30. Nguyen Ho PT, Vernooij MW, Voortman T, Rodriguez-Ayllon M, Neitzel J. Objective physical activity and Alzheimer’s disease burden in the population-based Rotterdam Study. Alzheimer’s & Dementia. 2025;21. 10.1002/alz.70655

31. Rodriguez-Ayllon M, Solis-Urra P, Arroyo-Ávila C, Álvarez-Ortega M, Molina-García P, Molina-Hidalgo C, et al. Physical activity and amyloid beta in middle-aged and older adults: A systematic review and meta-analysis. J Sport Health Sci. 2024;13:133–44. 10.1016/j.jshs.2023.08.001

32. Kimura N, Aso Y, Yabuuchi K, Ishibashi M, Hori D, Sasaki Y, et al. Association of Modifiable Lifestyle Factors With Cortical Amyloid Burden and Cerebral Glucose Metabolism in Older Adults With Mild Cognitive Impairment. JAMA Netw Open. 2020;3:e205719. 10.1001/jamanetworkopen.2020.5719

33. Kim SA, Shin D, Ham H, Kim Y, Gu Y, Kim HJ, et al. Physical Activity, Alzheimer Plasma Biomarkers, and Cognition. JAMA Netw Open. 2025;8:e250096. 10.1001/jamanetworkopen.2025.0096

34. Raffin J, Blennow K, Rolland Y, Cantet C, Guyonnet S, Vellas B, et al. Associations between moderate-to-vigorous physical activity, p-tau181, and cognition in healthy older adults with memory complaints: a secondary analysis from the MAPT. Lancet Healthy Longev. 2025;6:100678. 10.1016/j.lanhl.2024.100678

35. Brown BM, Rainey-Smith SR, Dore V, Peiffer JJ, Burnham SC, Laws SM, et al. Self-Reported Physical Activity is Associated with Tau Burden Measured by Positron Emission Tomography. Journal of Alzheimer’s Disease. 2018;63:1299–305. 10.3233/JAD-170998

36. Yau W-YW, Kirn DR, Rabin JS, Properzi MJ, Schultz AP, Shirzadi Z, et al. Physical activity as a modifiable risk factor in preclinical Alzheimer’s disease. Nat Med. 2025; 10.1038/s41591-025-03955-6

37. M. Tucker A, Stern Y. Cognitive Reserve in Aging. Curr Alzheimer Res [Internet]. 2011;8:354–60. 10.2174/156720511795745320

38. Song S, Stern Y, Gu Y. Modifiable lifestyle factors and cognitive reserve: A systematic review of current evidence. Ageing Res Rev [Internet]. 2022;74:101551. 10.1016/j.arr.2021.101551

39. Casaletto K, Ramos-Miguel A, VandeBunte A, Memel M, Buchman A, Bennett D, et al. Late-life physical activity relates to brain tissue synaptic integrity markers in older adults. Alzheimer’s & Dementia [Internet]. 2022;18:2023–35. 10.1002/alz.12530

40. Casaletto KB, Rentería MA, Pa J, Tom SE, Harrati A, Armstrong NM, et al. Late-Life Physical and Cognitive Activities Independently Contribute to Brain and Cognitive Resilience. Journal of Alzheimer’s Disease [Internet]. 2020;74:363–76. 10.3233/JAD-191114

41. Rabin JS, Klein H, Kirn DR, Schultz AP, Yang H-S, Hampton O, et al. Associations of Physical Activity and β-Amyloid With Longitudinal Cognition and Neurodegeneration in Clinically Normal Older Adults. JAMA Neurol. 2019;76:1203. 10.1001/jamaneurol.2019.1879

42. Dougherty RJ, Soldan A, Pettigrew C, Greenberg B, Spira AP, Moghekar A, et al. Physical activity modifies associations between cerebrospinal fluid tau measures and executive function. Alzheimer’s & Dementia: Translational Research & Clinical Interventions. 2025;11. 10.1002/trc2.70085

43. Anatürk M, Kaufmann T, Cole JH, Suri S, Griffanti L, Zsoldos E, et al. Prediction of brain age and cognitive age: Quantifying brain and cognitive maintenance in aging. Hum Brain Mapp [Internet]. 2021;42:1626–40. 10.1002/hbm.25316

44. Buchman AS, Yu L, Wilson RS, Lim A, Dawe RJ, Gaiteri C, et al. Physical activity, common brain pathologies, and cognition in community-dwelling older adults. Neurology [Internet]. 2019;92. 10.1212/WNL.0000000000006954

45. Yao T, Sweeney E, Nagorski J, Shulman JM, Allen GI. Quantifying cognitive resilience in Alzheimer’s Disease: The Alzheimer’s Disease Cognitive Resilience Score. Ginsberg SD, editor. PLoS One [Internet]. 2020;15:e0241707. 10.1371/journal.pone.0241707

46. Hamer M, Chida Y. Physical activity and risk of neurodegenerative disease: a systematic review of prospective evidence. Psychol Med [Internet]. 2009;39:3–11. 10.1017/S0033291708003681

47. Aghjayan SL, Lesnovskaya A, Esteban-Cornejo I, Peven JC, Stillman CM, Erickson KI. Aerobic exercise, cardiorespiratory fitness, and the human hippocampus. Hippocampus [Internet]. 2021;31:817–44. 10.1002/hipo.23337

48. Boots EA, Schultz SA, Oh JM, Larson J, Edwards D, Cook D, et al. Cardiorespiratory fitness is associated with brain structure, cognition, and mood in a middle-aged cohort at risk for Alzheimer’s disease. Brain Imaging Behav [Internet]. 2015;9:639–49. 10.1007/s11682-014-9325-9

49. Cole RC, Hazeltine E, Weng TB, Wharff C, DuBose LE, Schmid P, et al. Cardiorespiratory fitness and hippocampal volume predict faster episodic associative learning in older adults. Hippocampus [Internet]. 2020;30:143–55. 10.1002/hipo.23151

50. Erickson KI, Prakash RS, Voss MW, Chaddock L, Hu L, Morris KS, et al. Aerobic fitness is associated with hippocampal volume in elderly humans. Hippocampus [Internet]. 2009;19:1030–9. 10.1002/hipo.20547

51. Stillman CM, Uyar F, Huang H, Grove GA, Watt JC, Wollam ME, et al. Cardiorespiratory fitness is associated with enhanced hippocampal functional connectivity in healthy young adults. Hippocampus [Internet]. 2018;28:239–47. 10.1002/hipo.22827

52. Choi SH, Bylykbashi E, Chatila ZK, Lee SW, Pulli B, Clemenson GD, et al. Combined adult neurogenesis and BDNF mimic exercise effects on cognition in an Alzheimer’s mouse model. Science (1979) [Internet]. 2018;361. 10.1126/science.aan8821

53. van Praag H, Shubert T, Zhao C, Gage FH. Exercise Enhances Learning and Hippocampal Neurogenesis in Aged Mice. The Journal of Neuroscience [Internet]. 2005;25:8680–5. 10.1523/JNEUROSCI.1731-05.2005

54. van Praag H, Kempermann G, Gage FH. Running increases cell proliferation and neurogenesis in the adult mouse dentate gyrus. Nat Neurosci [Internet]. 1999;2:266–70. 10.1038/6368

55. Vaynman S, Ying Z, Gomez-Pinilla F. Hippocampal BDNF mediates the efficacy of exercise on synaptic plasticity and cognition. European Journal of Neuroscience [Internet]. 2004;20:2580–90. 10.1111/j.1460-9568.2004.03720.x

56. Vivar C, Potter MC, van Praag H. All About Running: Synaptic Plasticity, Growth Factors and Adult Hippocampal Neurogenesis. 2013. p. 189–210. 10.1007/7854_2012_220

57. Boa Sorte Silva NC, Barha CK, Erickson KI, Kramer AF, Liu-Ambrose T. Physical exercise, cognition, and brain health in aging. Trends Neurosci [Internet]. 2024;47:402–17. 10.1016/j.tins.2024.04.004

58. Leal LG, Lopes MA, Batista ML. Physical Exercise-Induced Myokines and Muscle-Adipose Tissue Crosstalk: A Review of Current Knowledge and the Implications for Health and Metabolic Diseases. Front Physiol [Internet]. 2018;9. 10.3389/fphys.2018.01307

59. Pedersen BK. Physical activity and muscle–brain crosstalk. Nat Rev Endocrinol [Internet]. 2019;15:383–92. 10.1038/s41574-019-0174-x

60. Pedersen BK, Åkerström TCA, Nielsen AR, Fischer CP. Role of myokines in exercise and metabolism. J Appl Physiol [Internet]. 2007;103:1093–8. 10.1152/japplphysiol.00080.2007

61. Wrann CD. FNDC5/Irisin – Their Role in the Nervous System and as a Mediator for Beneficial Effects of Exercise on the Brain. van Praag H, Christie B, editors. Brain Plasticity [Internet]. 2015;1:55–61. 10.3233/BPL-150019

62. Engeroff T, Füzéki E, Vogt L, Fleckenstein J, Schwarz S, Matura S, et al. Is Objectively Assessed Sedentary Behavior, Physical Activity and Cardiorespiratory Fitness Linked to Brain Plasticity Outcomes in Old Age? Neuroscience [Internet]. 2018;388:384–92. 10.1016/j.neuroscience.2018.07.050

63. Erickson KI, Voss MW, Prakash RS, Basak C, Szabo A, Chaddock L, et al. Exercise training increases size of hippocampus and improves memory. Proceedings of the National Academy of Sciences [Internet]. 2011;108:3017–22. 10.1073/pnas.1015950108

64. Gökçe E, Gün N. The Relationship Between Exercise, Cathepsin B, and Cognitive Functions: Systematic Review. Percept Mot Skills. 2023;130:1366–85. 10.1177/00315125231176980

65. Moon HY, Becke A, Berron D, Becker B, Sah N, Benoni G, et al. Running-Induced Systemic Cathepsin B Secretion Is Associated with Memory Function. Cell Metab [Internet]. 2016;24:332–40. 10.1016/j.cmet.2016.05.025

66. Gaitán JM, Moon HY, Stremlau M, Dubal DB, Cook DB, Okonkwo OC, et al. Effects of Aerobic Exercise Training on Systemic Biomarkers and Cognition in Late Middle-Aged Adults at Risk for Alzheimer’s Disease. Front Endocrinol (Lausanne) [Internet]. 2021;12. 10.3389/fendo.2021.660181

67. De la Rosa A, Solana E, Corpas R, Bartrés-Faz D, Pallàs M, Vina J, et al. Long-term exercise training improves memory in middle-aged men and modulates peripheral levels of BDNF and Cathepsin B. Sci Rep [Internet]. 2019;9:3337. 10.1038/s41598-019-40040-8

68. Vandersmissen J, Dewachter I, Cuypers K, Hansen D. The Impact of Exercise Training on the Brain and Cognition in Type 2 Diabetes, and its Physiological Mediators: A Systematic Review. Sports Med Open. 2025;11:42. 10.1186/s40798-025-00836-7

69. Morland C, Andersson KA, Haugen ØP, Hadzic A, Kleppa L, Gille A, et al. Exercise induces cerebral VEGF and angiogenesis via the lactate receptor HCAR1. Nat Commun. 2017;8:15557. 10.1038/ncomms15557

70. Leckie RL, Oberlin LE, Voss MW, Prakash RS, Szabo-Reed A, Chaddock-Heyman L, et al. BDNF mediates improvements in executive function following a 1-year exercise intervention. Front Hum Neurosci [Internet]. 2014;8:985. 10.3389/fnhum.2014.00985

71. Althubaiti A. Information bias in health research: definition, pitfalls, and adjustment methods. J Multidiscip Healthc [Internet]. 2016;211. 10.2147/JMDH.S104807

72. Caspersen CJ, Powell KE, Christenson GM. Physical activity, exercise, and physical fitness: definitions and distinctions for health-related research. Public Health Rep [Internet]. 1985;100:126–31. http://www.ncbi.nlm.nih.gov/pubmed/3920711

73. Arosio B, Calvani R, Ferri E, Coelho-Junior HJ, Carandina A, Campanelli F, et al. Sarcopenia and Cognitive Decline in Older Adults: Targeting the Muscle–Brain Axis. Nutrients [Internet]. 2023;15:1853. 10.3390/nu15081853

74. Cruz-Jentoft AJ, Bahat G, Bauer J, Boirie Y, Bruyère O, Cederholm T, et al. Sarcopenia: revised European consensus on definition and diagnosis. Age Ageing [Internet]. 2019;48:16–31. 10.1093/ageing/afy169

75. Behrenbruch N, Schwarck S, Schumann-Werner B, Molloy EN, Garcia-Garcia B, Hochkeppler A, et al. A physically and mentally active lifestyle relates to younger brain and cognitive age. Geroscience [Internet]. 2025; 10.1007/s11357-025-01764-w

76. Schmid NS, Ehrensperger MM, Berres M, Beck IR, Monsch AU. The Extension of the German CERAD Neuropsychological Assessment Battery with Tests Assessing Subcortical, Executive and Frontal Functions Improves Accuracy in Dementia Diagnosis. Dement Geriatr Cogn Dis Extra [Internet]. 2014;4:322–34. 10.1159/000357774

77. Jak AJ, Bondi MW, Delano-Wood L, Wierenga C, Corey-Bloom J, Salmon DP, et al. Quantification of Five Neuropsychological Approaches to Defining Mild Cognitive Impairment. The American Journal of Geriatric Psychiatry [Internet]. 2009;17:368–75. 10.1097/JGP.0b013e31819431d5

78. Helmstaedter C DH. The verbal learning and retention test. A useful and differentiated tool in evaluating verbal memory performance. Schweiz Arch Neurol Psychiatr. 1990;141:21–30.

79. Wechsler D. Wechsler Memory Scale-Revised: Manual. Psychological Corporation. 1987;

80. Buschke H. Cued recall in amnesia. J Clin Neuropsychol. 1984;6:433–40.

81. Meyers JE MK. Rey complex figure test under four different administration procedures. Clin Neuropsychol. 1995;9:63–7.

82. Smith A. Symbol digit modalities test. PsycTESTS Dataset. 2016;

83. Aschenbrenner S TOLK. Regensburger Wortflüssigkeits-Test. RWT Verlag für Psychologie: Hogrefe. 2000;

84. Papp K V., Rentz DM, Orlovsky I, Sperling RA, Mormino EC. Optimizing the preclinical Alzheimer’s cognitive composite with semantic processing: The PACC5. Alzheimer’s & Dementia: Translational Research & Clinical Interventions [Internet]. 2017;3:668–77. 10.1016/j.trci.2017.10.004

85. Xie L, Wisse LEM, Pluta J, de Flores R, Piskin V, Manjón J V., et al. Automated segmentation of medial temporal lobe subregions on in vivo T1-weighted MRI in early stages of Alzheimer’s disease. Hum Brain Mapp [Internet]. 2019;40:3431–51. 10.1002/hbm.24607

86. Boccardi M, Bocchetta M, Apostolova LG, Barnes J, Bartzokis G, Corbetta G, et al. Delphi definition of the EADC-ADNI Harmonized Protocol for hippocampal segmentation on magnetic resonance. Alzheimer’s & Dementia [Internet]. 2015;11:126–38. 10.1016/j.jalz.2014.02.009

87. Frisoni GB, Jack CR, Bocchetta M, Bauer C, Frederiksen KS, Liu Y, et al. The EADC-ADNI Harmonized Protocol for manual hippocampal segmentation on magnetic resonance: Evidence of validity. Alzheimer’s & Dementia [Internet]. 2015;11:111–25. 10.1016/j.jalz.2014.05.1756

88. Valdés Hernández M del C, Duarte Coello R, Xu W, Bernal J, Cheng Y, Ballerini L, et al. Influence of threshold selection and image sequence in in-vivo segmentation of enlarged perivascular spaces. J Neurosci Methods [Internet]. 2024;403:110037. 10.1016/j.jneumeth.2023.110037

89. Raz N, Daugherty AM, Bender AR, Dahle CL, Land S. Volume of the hippocampal subfields in healthy adults: differential associations with age and a pro-inflammatory genetic variant. Brain Struct Funct [Internet]. 2015;220:2663–74. 10.1007/s00429-014-0817-6

90. Potter GM, Chappell FM, Morris Z, Wardlaw JM. Cerebral Perivascular Spaces Visible on Magnetic Resonance Imaging: Development of a Qualitative Rating Scale and its Observer Reliability. Cerebrovascular Diseases [Internet]. 2015;39:224–31. 10.1159/000375153

91. Isensee F, Schell M, Pflueger I, Brugnara G, Bonekamp D, Neuberger U, et al. Automated brain extraction of multisequence MRI using artificial neural networks. Hum Brain Mapp [Internet]. 2019;40:4952–64. 10.1002/hbm.24750

92. Joshi AA, Choi S, Liu Y, Chong M, Sonkar G, Gonzalez-Martinez J, et al. A hybrid high-resolution anatomical MRI atlas with sub-parcellation of cortical gyri using resting fMRI. J Neurosci Methods [Internet]. 2022;374:109566. 10.1016/j.jneumeth.2022.109566

93. Mormino EC, Toueg TN, Azevedo C, Castillo JB, Guo W, Nadiadwala A, et al. Tau PET imaging with 18F-PI-2620 in aging and neurodegenerative diseases. Eur J Nucl Med Mol Imaging [Internet]. 2021;48:2233–44. 10.1007/s00259-020-04923-7

94. Rullmann M, Brendel M, Schroeter ML, Saur D, Levin J, Perneczky RG, et al. Multicenter 18F-PI-2620 PET for In Vivo Braak Staging of Tau Pathology in Alzheimer’s Disease. Biomolecules [Internet]. 2022;12:458. 10.3390/biom12030458

95. Kroth H, Oden F, Molette J, Schieferstein H, Capotosti F, Mueller A, et al. Discovery and preclinical characterization of [18F]PI-2620, a next-generation tau PET tracer for the assessment of tau pathology in Alzheimer’s disease and other tauopathies. Eur J Nucl Med Mol Imaging. 2019;46:2178–89. 10.1007/s00259-019-04397-2

96. Ichise M, Liow J-S, Lu J-Q, Takano A, Model K, Toyama H, et al. Linearized Reference Tissue Parametric Imaging Methods: Application to [11 C]DASB Positron Emission Tomography Studies of the Serotonin Transporter in Human Brain. Journal of Cerebral Blood Flow & Metabolism. 2003;23:1096–112. 10.1097/01.WCB.0000085441.37552.CA

97. López-González FJ, Paredes-Pacheco J, Thurnhofer-Hemsi K, Rossi C, Enciso M, Toro-Flores D, et al. QModeling: a Multiplatform, Easy-to-Use and Open-Source Toolbox for PET Kinetic Analysis. Neuroinformatics. 2019;17:103–14. 10.1007/s12021-018-9384-y

98. Brendel M, Barthel H, van Eimeren T, Marek K, Beyer L, Song M, et al. Assessment of 18 F-PI-2620 as a Biomarker in Progressive Supranuclear Palsy. JAMA Neurol [Internet]. 2020;77:1408. 10.1001/jamaneurol.2020.2526

99. Want A, Morgan JE, Barde Y-A. Brain-derived neurotrophic factor measurements in mouse serum and plasma using a sensitive and specific enzyme-linked immunosorbent assay. Sci Rep [Internet]. 2023;13:7740. 10.1038/s41598-023-34262-0

100. Morgado B, Klafki H-W, Bauer C, Waniek K, Esselmann H, Wirths O, et al. Assessment of immunoprecipitation with subsequent immunoassays for the blood-based diagnosis of Alzheimer’s disease. Eur Arch Psychiatry Clin Neurosci [Internet]. 2024; 10.1007/s00406-023-01751-2

101. Wickham H, Averick M, Bryan J, Chang W, McGowan L, François R, et al. Welcome to the Tidyverse. J Open Source Softw [Internet]. 2019;4:1686. 10.21105/joss.01686

102. Wei T, Simko V. corrplot: Visualization of a Correlation Matrix [Internet]. CRAN: Contributed Packages. 2010. 10.32614/CRAN.package.corrplot

103. den Heijer T, van der Lijn F, Koudstaal PJ, Hofman A, van der Lugt A, Krestin GP, et al. A 10-year follow-up of hippocampal volume on magnetic resonance imaging in early dementia and cognitive decline. Brain [Internet]. 2010;133:1163–72. 10.1093/brain/awq048

104. Meng X, D’Arcy C. Education and Dementia in the Context of the Cognitive Reserve Hypothesis: A Systematic Review with Meta-Analyses and Qualitative Analyses. Laks J, editor. PLoS One [Internet]. 2012;7:e38268. 10.1371/journal.pone.0038268

105. Tingley D, Yamamoto T, Hirose K, Keele L, Imai K. mediation: R Package for Causal Mediation Analysis. J Stat Softw [Internet]. 2014;59. 10.18637/jss.v059.i05

106. Preacher KJ, Hayes AF. Asymptotic and resampling strategies for assessing and comparing indirect effects in multiple mediator models. Behav Res Methods [Internet]. 2008;40:879–91. 10.3758/BRM.40.3.879

107. Preacher KJ, Hayes AF. SPSS and SAS procedures for estimating indirect effects in simple mediation models. Behavior Research Methods, Instruments, & Computers [Internet]. 2004;36:717–31. 10.3758/BF03206553

108. Braun M, Iliff JJ. The impact of neurovascular, blood-brain barrier, and glymphatic dysfunction in neurodegenerative and metabolic diseases. 2020. p. 413–36. 10.1016/bs.irn.2020.02.006

109. Greenberg SM, Bacskai BJ, Hernandez-Guillamon M, Pruzin J, Sperling R, van Veluw SJ. Cerebral amyloid angiopathy and Alzheimer disease — one peptide, two pathways. Nat Rev Neurol [Internet]. 2020;16:30–42. 10.1038/s41582-019-0281-2

110. Rasmussen MK, Mestre H, Nedergaard M. The glymphatic pathway in neurological disorders. Lancet Neurol [Internet]. 2018;17:1016–24. 10.1016/S1474-4422(18)30318-1

111. Tarasoff-Conway JM, Carare RO, Osorio RS, Glodzik L, Butler T, Fieremans E, et al. Clearance systems in the brain—implications for Alzheimer disease. Nat Rev Neurol [Internet]. 2015;11:457–70. 10.1038/nrneurol.2015.119

112. Wardlaw JM, Benveniste H, Nedergaard M, Zlokovic B V., Mestre H, Lee H, et al. Perivascular spaces in the brain: anatomy, physiology and pathology. Nat Rev Neurol [Internet]. 2020;16:137–53. 10.1038/s41582-020-0312-z

113. Francis F, Ballerini L, Wardlaw JM. Perivascular spaces and their associations with risk factors, clinical disorders and neuroimaging features: A systematic review and meta-analysis. International Journal of Stroke [Internet]. 2019;14:359–71. 10.1177/1747493019830321

114. He X, Liu D, Zhang Q, Liang F, Dai G, Zeng J, et al. Voluntary Exercise Promotes Glymphatic Clearance of Amyloid Beta and Reduces the Activation of Astrocytes and Microglia in Aged Mice. Front Mol Neurosci [Internet]. 2017;10. 10.3389/fnmol.2017.00144

115. Bliss ES, Wong RH, Howe PR, Mills DE. Benefits of exercise training on cerebrovascular and cognitive function in ageing. J Cereb Blood Flow Metab [Internet]. 2021;41:447–70. 10.1177/0271678X20957807

116. He X-F, Liu D-X, Zhang Q, Liang F-Y, Dai G-Y, Zeng J-S, et al. Voluntary Exercise Promotes Glymphatic Clearance of Amyloid Beta and Reduces the Activation of Astrocytes and Microglia in Aged Mice. Front Mol Neurosci [Internet]. 2017;10:144. 10.3389/fnmol.2017.00144

117. Yang Y, Wang M, Luan M, Song X, Wang Y, Xu L, et al. Enlarged Perivascular Spaces and Age-Related Clinical Diseases. Clin Interv Aging [Internet]. 2023;18:855–67. 10.2147/CIA.S404908

118. Nguyen Ho PT, Vernooij MW, Voortman T, Rodriguez-Ayllon M, Neitzel J. Objective physical activity and Alzheimer’s disease burden in the population-based Rotterdam Study. Alzheimer’s & Dementia. 2025;21. 10.1002/alz.70655

119. Pereira JB, Janelidze S, Smith R, Mattsson-Carlgren N, Palmqvist S, Teunissen CE, et al. Plasma GFAP is an early marker of amyloid-β but not tau pathology in Alzheimer’s disease. Brain. 2021;144:3505–16. 10.1093/brain/awab223

120. Saur L, Baptista PPA, de Senna PN, Paim MF, Nascimento P do, Ilha J, et al. Physical exercise increases GFAP expression and induces morphological changes in hippocampal astrocytes. Brain Struct Funct. 2014;219:293–302. 10.1007/s00429-012-0500-8

121. VandeBunte AM, Lee SY, Paolillo EW, Rojas JC, Chan B, Lago AL, et al. Physical Activity Relates to Lower Astrocytic Activation and Axonal Breakdown in Clinically Normal Older Adults. Alzheimer’s & Dementia. 2022;18. 10.1002/alz.063455

122. Raffin J. Does Physical Exercise Modify the Pathophysiology of Alzheimer’s Disease in Older Persons? The Journal of Aging Research & Lifestyle. 2024;13:77–81. 10.14283/jarlife.2024.11

123. Roccati E, Collins JM, Bindoff AD, Alty JE, Bartlett L, King AE, et al. Modifiable risk factors for dementia, cognition, and plasma phosphorylated tau 181 in a large-scale cohort of Australian older adults. Neurobiol Aging. 2023;131:106–14. 10.1016/j.neurobiolaging.2023.06.018

124. Roccati E, Bindoff AD, Collins JM, Eastgate J, Borchard J, Alty J, et al. Modifiable dementia risk factors and AT(N) biomarkers: findings from the EPAD cohort. Front Aging Neurosci. 2024;16. 10.3389/fnagi.2024.1346214

125. Zhong S, Zhao B, Ma Y-H, Sun Y, Zhao Y-L, Liu W-H, et al. Associations of Physical Activity with Alzheimer’s Disease Pathologies and Cognition: The CABLE Study. Journal of Alzheimer’s Disease. 2022;89:483–92. 10.3233/JAD-220389

126. Law LL, Rol RN, Schultz SA, Dougherty RJ, Edwards DF, Koscik RL, et al. Moderate intensity physical activity associates with CSF biomarkers in a cohort at risk for Alzheimer’s disease. Alzheimer’s & Dementia: Diagnosis, Assessment & Disease Monitoring. 2018;10:188–95. 10.1016/j.dadm.2018.01.001

127. Hou X-H, Xu W, Bi Y-L, Shen X-N, Ma Y-H, Dong Q, et al. Associations of healthy lifestyles with cerebrospinal fluid biomarkers of Alzheimer’s disease pathology in cognitively intact older adults: the CABLE study. Alzheimers Res Ther. 2021;13:81. 10.1186/s13195-021-00822-7

128. Braak H, Braak E. Neuropathological stageing of Alzheimer-related changes. Acta Neuropathol [Internet]. 1991;82:239–59. 10.1007/BF00308809

129. Braak H, Braak E. Frequency of Stages of Alzheimer-Related Lesions in Different Age Categories. Neurobiol Aging [Internet]. 1997;18:351–7. 10.1016/S0197-4580(97)00056-0

130. Crary JF, Trojanowski JQ, Schneider JA, Abisambra JF, Abner EL, Alafuzoff I, et al. Primary age-related tauopathy (PART): a common pathology associated with human aging. Acta Neuropathol [Internet]. 2014;128:755–66. 10.1007/s00401-014-1349-0

131. Jagust W. Imaging the evolution and pathophysiology of Alzheimer disease. Nat Rev Neurosci [Internet]. 2018;19:687–700. 10.1038/s41583-018-0067-3

132. Masters CL, Bateman R, Blennow K, Rowe CC, Sperling RA, Cummings JL. Alzheimer’s disease. Nat Rev Dis Primers [Internet]. 2015;1:15056. 10.1038/nrdp.2015.56

133. Braak H, Alafuzoff I, Arzberger T, Kretzschmar H, Del Tredici K. Staging of Alzheimer disease-associated neurofibrillary pathology using paraffin sections and immunocytochemistry. Acta Neuropathol [Internet]. 2006;112:389–404. 10.1007/s00401-006-0127-z

134. Maass A, Lockhart SN, Harrison TM, Bell RK, Mellinger T, Swinnerton K, et al. Entorhinal Tau Pathology, Episodic Memory Decline, and Neurodegeneration in Aging. The Journal of Neuroscience [Internet]. 2018;38:530–43. 10.1523/JNEUROSCI.2028-17.2017

135. Groot C, Doré V, Robertson J, Burnham SC, Savage G, Ossenkoppele R, et al. Mesial temporal tau is related to worse cognitive performance and greater neocortical tau load in amyloid-β–negative cognitively normal individuals. Neurobiol Aging. 2021;97:41–8. 10.1016/j.neurobiolaging.2020.09.017

136. Ziontz J, Bilgel M, Shafer AT, Moghekar A, Elkins W, Helphrey J, et al. Tau pathology in cognitively normal older adults. Alzheimer’s & Dementia: Diagnosis, Assessment & Disease Monitoring. 2019;11:637–45. 10.1016/j.dadm.2019.07.007

137. Sperling RA, Mormino EC, Schultz AP, Betensky RA, Papp K V., Amariglio RE, et al. The impact of amyloid-beta and tau on prospective cognitive decline in older individuals. Ann Neurol. 2019;85:181–93. 10.1002/ana.25395

138. Lowe VJ, Bruinsma TJ, Wiste HJ, Min H-K, Weigand SD, Fang P, et al. Cross-sectional associations of tau-PET signal with cognition in cognitively unimpaired adults. Neurology. 2019;93. 10.1212/WNL.0000000000007728

139. Dougherty RJ, Soldan A, Pettigrew C, Greenberg B, Spira AP, Moghekar A, et al. Physical activity modifies associations between cerebrospinal fluid tau measures and executive function. Alzheimer’s & Dementia: Translational Research & Clinical Interventions. 2025;11. 10.1002/trc2.70085

140. Stillman CM, Esteban-Cornejo I, Brown B, Bender CM, Erickson KI. Effects of Exercise on Brain and Cognition Across Age Groups and Health States. Trends Neurosci [Internet]. 2020;43:533–43. 10.1016/j.tins.2020.04.010

141. Duzel E, van Praag H, Sendtner M. Can physical exercise in old age improve memory and hippocampal function? Brain [Internet]. 2016;139:662–73. 10.1093/brain/awv407

142. van Praag H, Christie BR, Sejnowski TJ, Gage FH. Running enhances neurogenesis, learning, and long-term potentiation in mice. Proceedings of the National Academy of Sciences [Internet]. 1999;96:13427–31. 10.1073/pnas.96.23.13427

143. Voss MW, Vivar C, Kramer AF, van Praag H. Bridging animal and human models of exercise-induced brain plasticity. Trends Cogn Sci [Internet]. 2013;17:525–44. 10.1016/j.tics.2013.08.001

144. Jodeiri Farshbaf M, Alviña K. Multiple Roles in Neuroprotection for the Exercise Derived Myokine Irisin. Front Aging Neurosci [Internet]. 2021;13. 10.3389/fnagi.2021.649929

145. Lu Y, Bu F-Q, Wang F, Liu L, Zhang S, Wang G, et al. Recent advances on the molecular mechanisms of exercise-induced improvements of cognitive dysfunction. Transl Neurodegener [Internet]. 2023;12:9. 10.1186/s40035-023-00341-5

146. Chaddock L, Erickson KI, Prakash RS, Kim JS, Voss MW, Vanpatter M, et al. A neuroimaging investigation of the association between aerobic fitness, hippocampal volume, and memory performance in preadolescent children. Brain Res [Internet]. 2010;1358:172–83. 10.1016/j.brainres.2010.08.049

147. Makizako H, Liu-Ambrose T, Shimada H, Doi T, Park H, Tsutsumimoto K, et al. Moderate-Intensity Physical Activity, Hippocampal Volume, and Memory in Older Adults With Mild Cognitive Impairment. J Gerontol A Biol Sci Med Sci [Internet]. 2015;70:480–6. 10.1093/gerona/glu136

148. Maass A, Düzel S, Brigadski T, Goerke M, Becke A, Sobieray U, et al. Relationships of peripheral IGF-1, VEGF and BDNF levels to exercise-related changes in memory, hippocampal perfusion and volumes in older adults. Neuroimage [Internet]. 2016;131:142–54. 10.1016/j.neuroimage.2015.10.084

