## Supplemental Material for "Physical fitness supports brain maintenance and cognitive reserve in cognitively normal aging: a cross-sectional examination"

In a secondary analysis, all components of the muscular capacity composite (TUG, ASMM, and maximal handgrip strength) were examined in relation to pathology markers. A significant negative association was observed between the Timed-Up-and-Go test (TUG, in seconds) and BG PVS volumes ($\beta$ = -0.283, SE = 0.094, $p_{adjusted}$ = .007; Fig S1A), indicating that better physical (walking) performance was associated with lower BG PVS volumes. To separate motor from executive contributions, the Trail Making Test ratio (TMT B/A), indexing executive control, was included as an additional covariate. TUG remained a significant predictor ($\beta$ = -0.28, SE = 0.09, p = .002), whereas TMT B/A showed no significant association with BG PVS volumes ($\beta$ = -0.03, SE = 0.06, p = .632). A negative association between TUG and WMH burden was also observed (p = .040), although this did not survive multiple comparison correction ($\beta$ = -0.18, SE = 0.09, $p_{adjusted}$ = .083).

By contrast, neither maximal handgrip strength nor ASMM showed significant associations with BG PVS (Fig S1B-C) or with any other pathology marker (all $p_{adjusted}$ > .05).

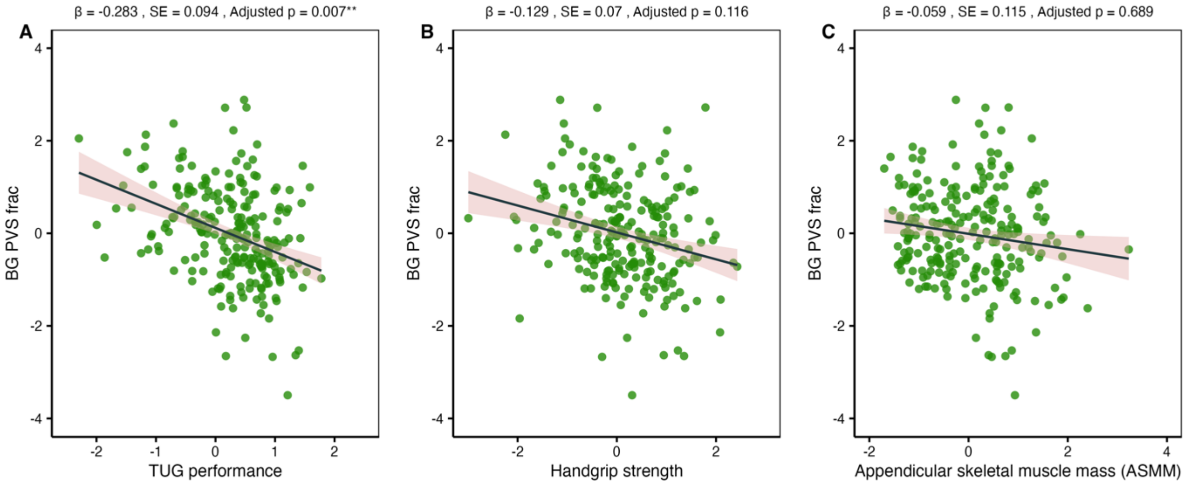


Fig. S1. Multiple linear regression plots illustrating the contribution of muscular fitness components to brain resistance. Effect of TUG (timed-up-and-go- est assessing physical performance; A), Handgrip strength (B), and ASMM (appendicular skeletal muscle mass; C) on BG PVS volumes (basal ganglia perivascular spaces, normalized for BG volume). The estimated regression line is displayed with its 95% confidence interval (light pink shaded area). β: regression slope (beta coefficient); SE: standard error; Adjusted p: FDR-corrected p-value (Benjamini–Hochberg procedure). * indicates Adjusted $p < .05$. All models were adjusted for age and sex.

Table S1: Descriptive statistic of physical fitness parameters

| Outcome | M | SD | Median | IQR |
|---|---|---|---|---|
| Handgrip | 30.87 | 9.50 | 29.83 | 14.75 |
| ASMM | 20.42 | 4.71 | 20.20 | 7.30 |
| TUG | 6.09 | 1.52 | 5.76 | 1.62 |
| VO2max | 22.18 | 5.92 | 21.30 | 7.52 |

Uncorrected Raw values are presented as mean (M), standard deviation (SD), median and interquartile range (IQR). Handgrip, maximal handgrip strength (average of three maximal voluntary contractions per hand); ASMM, appendicular skeletal muscle mass (in kg); TUG, timed-up-and-go-test (in seconds); $VO_2max$, maximal oxygen consumption (in ml/min/kg).

Table S2: Descriptive statistic of demographic and pathology parameters

| Outcome | M | SD | Median | IQR |
|---|---|---|---|---|
| Age | 72.77 | 7.95 | 71.00 | 15.00 |
| Education Years | 14.93 | 2.38 | 15.00 | 3.00 |
| PACC5 | −0.01 | 0.65 | 0.05 | 0.84 |
| VLMT | 10.33 | 2.96 | 10.00 | 5.00 |
| ROCFT | 17.95 | 6.10 | 18.00 | 8.00 |
| BG PVS | 0.17 | 0.12 | 0.14 | 0.11 |
| CSO PVS | 0.81 | 0.64 | 0.67 | 0.75 |
| WMH | 4.65 | 7.01 | 1.76 | 4.29 |
| MTL DVR | 0.93 | 0.06 | 0.93 | 0.09 |
| ptau 217 | 0.13 | 0.10 | 0.10 | 0.05 |
| Aβ42/40 | 0.09 | 0.01 | 0.09 | 0.01 |

Uncorrected Raw values are presented as mean (M), standard deviation (SD), median and interquartile range (IQR). Age and Education years in years; CERAD, Consortium to establish a Registry for Alzheimer's Disease Plus version (z-composite); PACC5, preclinical Alzheimer's composite (z-composite); VLMT, German adaptation of Rey's Auditory Verbal Learning Test (delayed verbal memory performance); ROCFT, Rey-Osterrieth Complex Figure Test (visuospatial delayed memory performance); BG PVS, total basal ganglia perivascular space volume (in mL); CSO PVS, total centrum semiovale perivascular space volume (in mL); WMH, total white matter hyperintensity volume (in mL); MTL DVR, , medial temporal lobe (MTL) tau load as distribution volume ratio (DVR); plasma pTau217 levels (in pg/ml) and the $A\beta_{1-42}$/ $A\beta_{1-40}$ ratio. All variables are shown as uncorrected raw values, except the PACC5 z-composite.

Table S3: Descriptive statistic of structural parameters

| Outcome | M | SD | Median | IQR |
|---|---|---|---|---|
| MTL thickness | 2.80 | 0.14 | 2.80 | 0.19 |
| bilat wHCV | 6935.03 | 597.86 | 6927.18 | 840.65 |
| bilat aHCV | 3546.81 | 424.38 | 3568.92 | 571.87 |
| bilat pHCV | 3388.21 | 330.91 | 3407.69 | 429.05 |
| left wHCV | 3437.58 | 313.96 | 3454.60 | 412.24 |
| left aHCV | 1732.86 | 217.34 | 1734.65 | 274.00 |
| left pHCV | 1704.72 | 176.39 | 1713.00 | 220.83 |
| right wHCV | 3497.44 | 317.66 | 3521.09 | 443.88 |
| right aHCV | 1813.95 | 239.79 | 1820.39 | 304.73 |
| right pHCV | 1683.49 | 170.56 | 1695.23 | 241.95 |

Uncorrected Raw values are presented as mean (M), standard deviation (SD), median and interquartile range (IQR). MTL thickness, medial temporal lobe thickness (in mm). Volumetric measures for the anterior hippocampus (aHCV), posterior hippocampus (pHCV), and total hippocampal volume (wHCV) are shown for both hemispheres (bilateral), as well as for the left and right sides. All volumes are expressed in $mm^3$ and corrected for total intracranial volume.
